# Evaluation of short-term multi-target respiratory forecasts over winter 2024-25 in England using sub-ensemble contribution analyses

**DOI:** 10.64898/2026.02.12.26346156

**Authors:** Jack Kennedy, William Ferguson, Owen Jones, Steven Riley, Thomas Ward, Maria L. Tang, Jonathon Mellor

## Abstract

**Background:** Epidemic forecasting research often assesses ensembles and their component models using probabilistic scoring rules. Quantifying how individual models affect ensemble performance is challenging, particularly across multiple targets and spatial scales.

**Methods:** We present Winter 2024-25 forecasts of Influenza and COVID-19 hospital admissions in England and conduct a retrospective simulation using the operational component models. Forecasts were scored using the per capita weighted interval score (pcWIS) for counts and the ranked probability score (RPS) for ordinal trend direction. We compared operational retrospective forecasts, used generalised additive models (GAMs) to estimate the expected change in score from the inclusion of a model in a sub-ensemble (an ensemble formed from a subset of available models), and used Pareto analysis to understand which sub-ensembles were Pareto-optimal across scoring rules.

**Results:** Nationally, there was a 47% improvement in Influenza pcWIS versus sub-ensembles. However, Influenza operational ensembles were on average 22% worse than sub-ensembles, when measured by RPS. For COVID-19, operational ensembles were 43% and 280% worse on average, than retrospective sub-ensembles by pcWIS and RPS, respectively. However, COVID-19 operational ensembles were on average 2% (pcWIS) and 13% (RPS) better than individual operational models. For influenza, operational ensembles were, on average, 58% (pcWIS) and 41% (RPS) better than individual models. The sub-ensemble simulation showed individual models influenced the ensembles during different epidemic phases. The Pareto analysis demonstrated that there can be a trade-off between relative direction and absolute count score optimisation.

**Interpretation:** Our analysis shows that UK Health Security Agency forecasts were well calibrated with observations and often had comparable performance to optimal ensembles. Our GAM and Pareto analyses inform model selection for future ensembles.

**Author Summary:** Forecasts of winter hospital pressures in England are an important tool for senior healthcare leaders. It is common practice to produce a forecasting ensemble, i.e. combine the predictions of multiple models to create a single, more accurate prediction. Forecasting teams should strive to produce the best forecast possible; one tool for this is retrospective evaluation over a forecasting season using proper scoring rules to assess performance. Our forecasts are constructed of two components, an epidemic trend direction estimate as well as forecast of hospital admission numbers. There are two main challenges we address. The first is understanding at which epidemic phase different ensemble contributions are most effective, the second is the joint optimisation of an ensemble for both trend direction and admission numbers forecast. We apply these methods to a variety of ensembles (sub-ensembles) based on our own modelling suite, and compare the sub-ensembles to our operational forecasts from the Winter 2024/25 season.

## Introduction

Respiratory pathogens, such as SARS-CoV-2 and Influenza, place substantial pressure on health systems in winter through emergency care visits, hospital admissions, and bed occupancy. The UK Health Security Agency (UKHSA) is responsible for prevention, and harm reduction of infectious diseases in England. UKHSA provides the National Health Service (NHS) and other public sector organisations in England with short-term forecasts (14 day forecast horizon) of hospital admissions due to COVID-19 and Influenza. UKHSA also provide a range of other real-time modelling products across pathogens such as RSV and norovirus [1] each winter, as well as during outbreak response [2, 3].

There is a rich history of using a combination of models, referred to as an ensemble, to forecast Influenza [4], COVID-19 [5], and other respiratory disease indicators [6]. Model ensembling helps to address limitations of individual models, producing a consensus estimate which can be less biased and is more robust than contributing models. While ensembles generally outperform individual models [7], not all ensemble methods are equally performant [8, 9]. The short-term ensemble forecasts produced by UKHSA in the winter 2023-24 season were shown to be useful, but with room for improvement [10]. In the past two winters, UKHSA has ensembled models via unweighted posterior stacking [11], which produces prediction samples from the ensemble. Many similar forecasting hubs and teams use quantile averaging techniques to construct ensembles; this is an accurate and practical approach [12] when stacking is impractical. For example, submitting a small collection of quantiles is computationally and operationally much simpler than submitting hundreds or thousands of posterior samples to a forecasting hub.

In real-time settings, the decision to include or exclude a model is typically guided by experience and judgement informed by performance and plausibility checks. This decision to include or not can be thought of as model weighting, where an excluded model within an ensemble is given zero weight. It is not straightforward to know in real time which models should be highly weighted at a given point in time [13, 14, 15]. However, knowledge of a model’s past performance (from previous seasons or more recently) may help modellers to understand which models are most appropriate at different epidemic phases [16]. Further, individual model performance does not translate in a straightforward way to a change in ensemble performance [17]. We therefore aim to develop methods to understand which models are most useful in ensembles during different epidemic phases.

For the winter 2024-25 season, UKHSA developed a suite of short-term forecast models for COVID-19, Influenza, RSV and norovirus. These forecasts were delivered widely within the health system in England. Users of UKHSA’s forecasts included:

- Senior officials and politicians in central government (DHSC) responsible for policy.
- National and local public health officials responsible for interventions, resourcing and surveillance.
- National healthcare system managers responsible for resilience, supply chains and system integration.
- Regional and local healthcare system managers responsible for capacity, bed and staff management in primary/secondary care.

In this paper we present an evaluation of the prospectively reported COVID-19 and Influenza hospital admission operational ensemble forecasts used for decision making, as well as a retrospective exploration of individual models’ contributions to sub-ensembles. Furthermore, we explore sub-ensembles, an ensemble formed from a subset of available models, to help draw more general conclusions about individual contributing models. We aim to understand how individual models contribute to forecast accuracy & precision by evaluating a range of possible sub-ensembles to compare their performance. We then explore how the individual model’s contribution changes over time and geography to help inform future seasons ensemble composition.

A common challenge in evaluating epidemic forecasts for practitioners in public health is the choice of evaluation scoring rule. Often a variety of rules are presented in research, but often only a single evaluation metric is used to guide model development. The choice of which evaluation metric to optimize against is challenging as different users have different needs. For example, a hospital capacity manager may be primarily interested in local absolute values of hospital admissions, whereas a national policy maker may care more about the national direction of an epidemic. Designing a model that is only good for one user type, but poor for another is unsatisfying and an area requiring further work. In this work we tackle the competing priorities in epidemic forecast evaluation using a Pareto front analysis.

## Methods

### Ethics statement

UKHSA have an exemption under regulation 3 of Sect. 251 of the National Health Service Act (2006) to allow identifiable patient information to be processed to diagnose, control, prevent, or recognise trends in, communicable diseases and other risks to public health.

### Forecast targets

Our forecasting targets are key metrics for winter hospital pressures, identified by working with healthcare operational managers. In the Winter 2024-25 season, UKHSA forecast new test-positive hospitalisations for COVID-19, Influenza, respiratory syncytial virus (RSV), and norovirus test positive cases. The basis of this paper is primarily to present and analyse forecasts of COVID-19 and Influenza hospital admissions with the most comprehensive operational ensembles. Both COVID-19 & Influenza operational ensembles contained more than three models, whereas the RSV operational ensemble contained only three models, and norovirus a single model. For this reason, we present and evaluate the RSV and norovirus forecasts in Supplementary Figures A - J. We exclude these pathogens from the retrospective sub-ensemble analysis due to having insufficient operational models developed this season.

Consistent and comprehensive data collection across hospitals in England is performed by the National Health Service (NHS). In secondary care settings, if an individual presents severe disease symptoms, a diagnostic test will be performed with results reported to UKHSA via the Second-Generation Surveillance System (SGSS). The data and forecasts are, for each date of a test, the number of unique inpatients with a new positive test taken within the past 24 hours. The data can be revised for up to 14 days following submission, however, in practice the extent of revisions is small shown in Supplementary Figure K. All forecasts are produced weekly, at daily granularity, with a 14-day forecast horizon.

The forecast targets discussed in this paper were produced at multiple spatial scales. Influenza and COVID-19 admissions were forecast at the level of integrated care boards (ICBs) and then aggregated to produce regional and national forecasts. An ICB is a sub-regional NHS body responsible for the management of local health services. ICBs are each nested within one of the seven NHS regions. A map illustrating NHS regions and ICBs is presented as Supplementary Figure L.

Our secondary forecast target approach is an epidemic trend direction assessment. We categorise an epidemic as one of “decreasing”, “stable”, or “increasing”, an ordinal classification. This is a simplification of our forecasts, but useful for many users. The trend direction for a given forecast day is estimated by first calculating the 7-day right aligned rolling average of forecast target and forecast sample value. The direction is then a percentage change between the rolling average target, and a rolling average forecast 14 days ahead (our forecast horizon). A change between target data and forecast value of ≥ 20% is considered an “increase”, while a downward change of ≥ 20% is a “decrease”. Changes less than 20% are considered “stable”. This procedure produces trend directions for each time step in the forecast horizon, up to the final forecast day. For forecast users we only show the final forecast trend direction as the most relevant overall indication of direction. This method has desirable qualities – we can use the approach across many pathogens without alteration and can deploy it at a range of spatial and population scales, which are more challenging with absolute changes.

We calculate the trend direction probability as the proportion of posterior samples that are between each threshold boundary, for example if 350 of 500 posterior sample predictions meet the “increase” definition, we have a probability of increase of 70%. The specific threshold of 20% was chosen following consultation with forecast users as a meaningful change in trend that maintained coherence with other surveillance metrics users were familiar with. Surveillance indicators within UKHSA and elsewhere are often classified using a 10% week-on-week change, which we extend to our 2-week horizon, which would be a 21% increase (1.1 × 1.1 =1.21). We approximate the 21% change to 20% to make the method clearer for users. We are currently considering how to extend this approach to even longer time horizons, but feedback from users will be the primary driver.

The individual models used operationally in the 2024-25 season are described in Table 1. Each GAM model is a permutation of previous work [18], with the ETS models also described in [10]. The GP growth rate, Mean GR and Median GR models were conceptually new to the English operational ensemble this year, with further details given in Supplementary Section B. Full implementation details are available in the supporting code https://github.com/jcken95/sub-ensemble-evaluation.

**Table 1.**
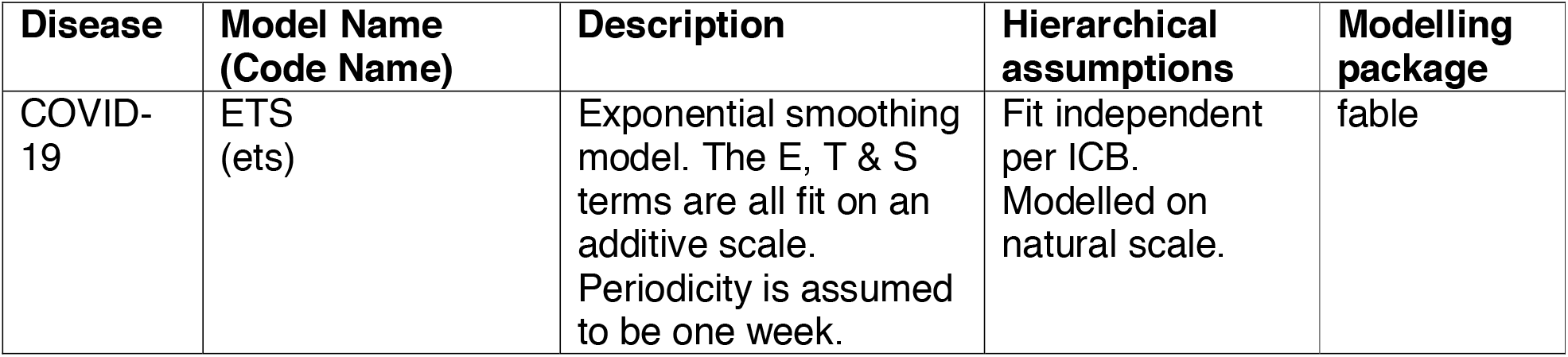
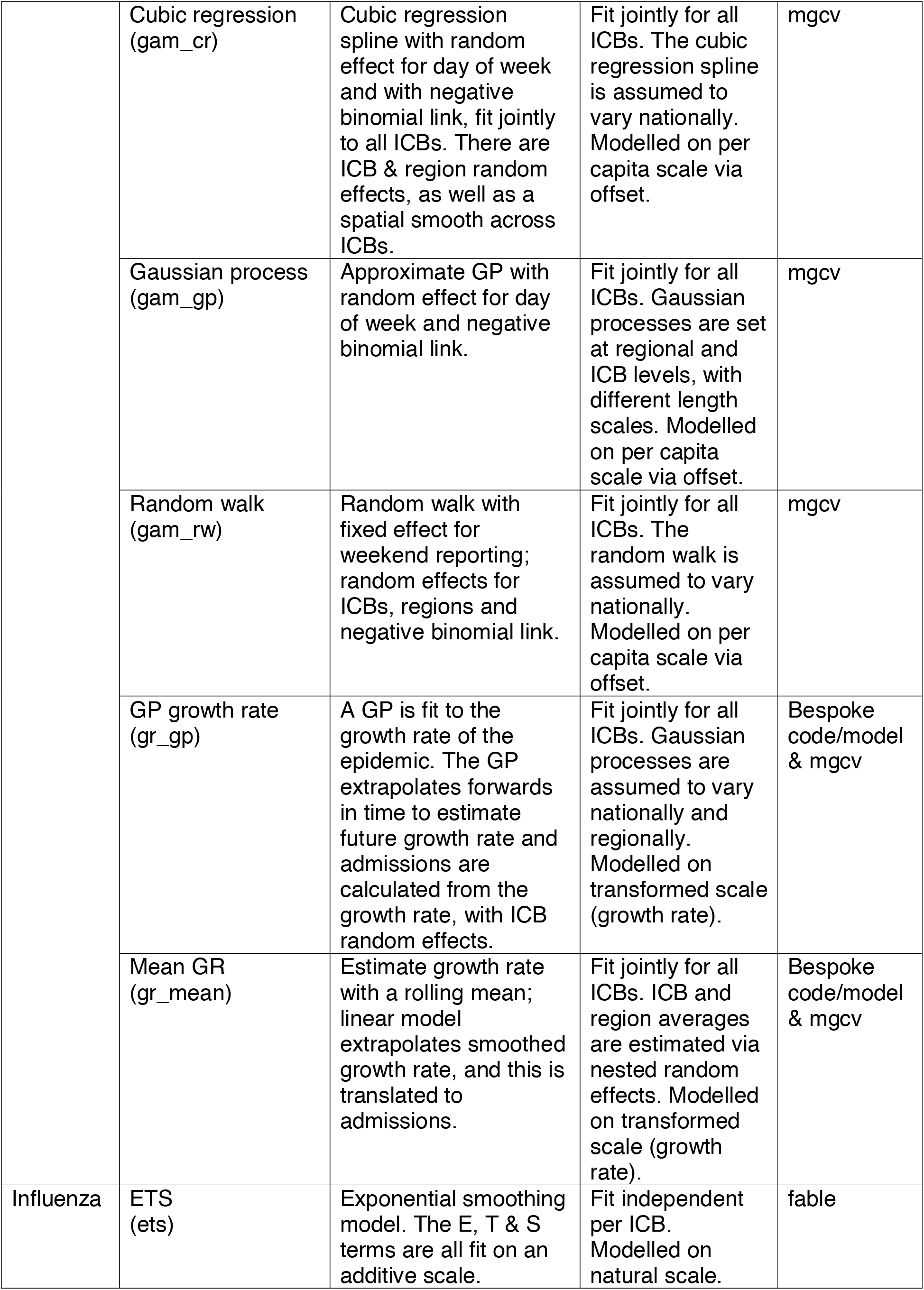
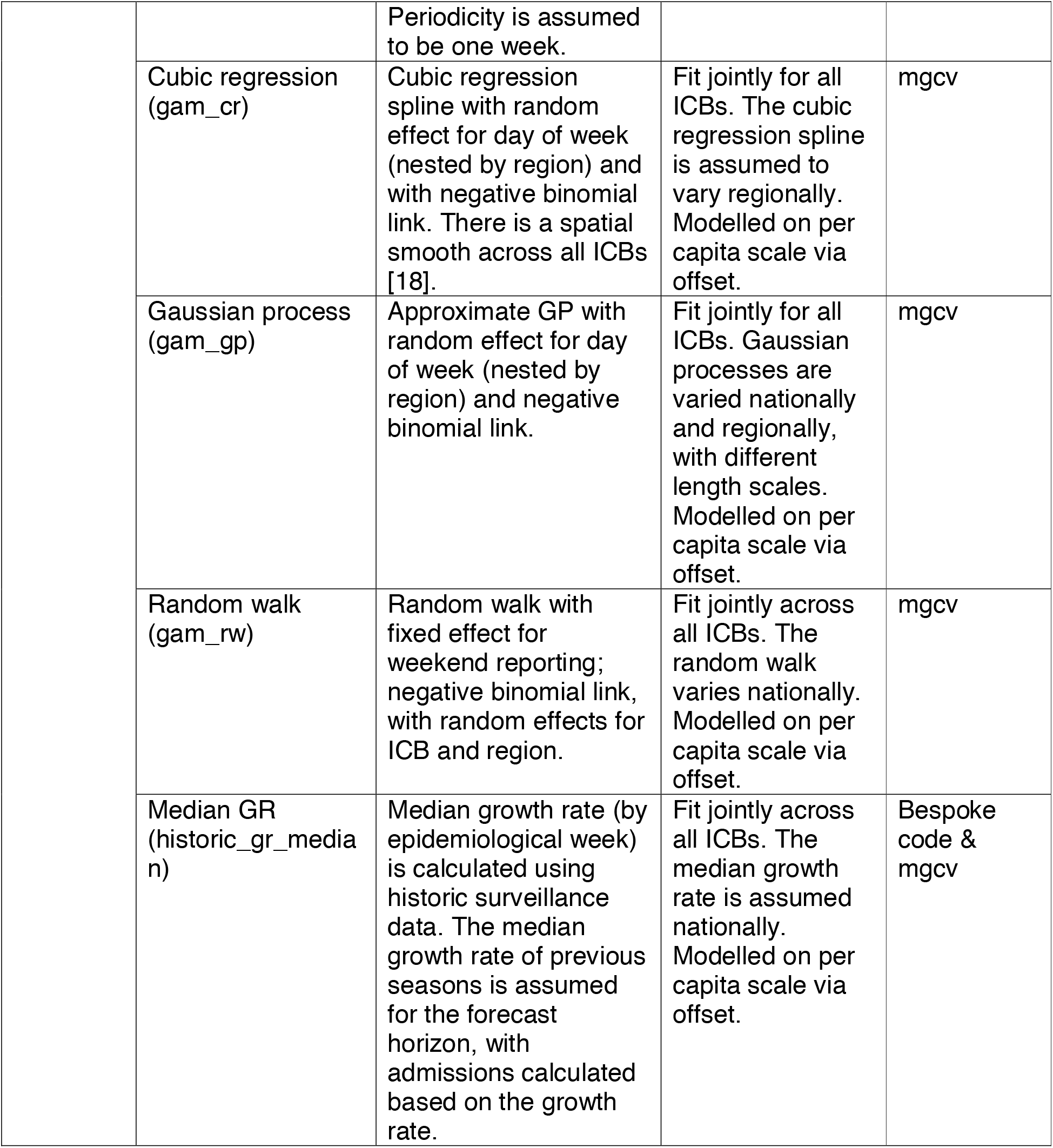
Name and description of each model used in during the winter 2024-25 season with supporting description and package name.

| Disease | Model Name<br>(Code Name) | Description | Hierarchical<br>assumptions | Modelling<br>package |
| --- | --- | --- | --- | --- |
| COVID-19 | ETS<br>(ets) | Exponential smoothing model. The E, T & S terms are all fit on an additive scale. Periodicity is assumed to be one week. | Fit independent per ICB. Modelled on natural scale. | fable |
|  | Cubic regression (gam_cr) | Cubic regression spline with random effect for day of week and with negative binomial link, fit jointly to all ICBs. There are ICB & region random effects, as well as a spatial smooth across ICBs. | Fit jointly for all ICBs. The cubic regression spline is assumed to vary nationally. Modelled on per capita scale via offset. | mgcv |
|  | Gaussian process (gam_gp) | Approximate GP with random effect for day of week and negative binomial link. | Fit jointly for all ICBs. Gaussian processes are set at regional and ICB levels, with different length scales. Modelled on per capita scale via offset. | mgcv |
|  | Random walk (gam_rw) | Random walk with fixed effect for weekend reporting; random effects for ICBs, regions and negative binomial link. | Fit jointly for all ICBs. The random walk is assumed to vary nationally. Modelled on per capita scale via offset. | mgcv |
|  | GP growth rate (gr_gp) | A GP is fit to the growth rate of the epidemic. The GP extrapolates forwards in time to estimate future growth rate and admissions are calculated from the growth rate, with ICB random effects. | Fit jointly for all ICBs. Gaussian processes are assumed to vary nationally and regionally. Modelled on transformed scale (growth rate). | Bespoke code/model & mgcv |
|  | Mean GR (gr_mean) | Estimate growth rate with a rolling mean; linear model extrapolates smoothed growth rate, and this is translated to admissions. | Fit jointly for all ICBs. ICB and region averages are estimated via nested random effects. Modelled on transformed scale (growth rate). | Bespoke code/model & mgcv |
| Influenza | ETS (ets) | Exponential smoothing model. The E, T & S terms are all fit on an additive scale. | Fit independent per ICB. Modelled on natural scale. | fable |
|  |  | Periodicity is assumed to be one week. |  |  |
|  | Cubic regression (gam_cr) | Cubic regression spline with random effect for day of week (nested by region) and with negative binomial link. There is a spatial smooth across all ICBs [18]. | Fit jointly for all ICBs. The cubic regression spline is assumed to vary regionally. Modelled on per capita scale via offset. | mgcv |
|  | Gaussian process (gam_gp) | Approximate GP with random effect for day of week (nested by region) and negative binomial link. | Fit jointly for all ICBs. Gaussian processes are varied nationally and regionally, with different length scales. Modelled on per capita scale via offset. | mgcv |
|  | Random walk (gam_rw) | Random walk with fixed effect for weekend reporting; negative binomial link, with random effects for ICB and region. | Fit jointly across all ICBs. The random walk varies nationally. Modelled on per capita scale via offset. | mgcv |
|  | Median GR (historic_gr_median) | Median growth rate (by epidemiological week) is calculated using historic surveillance data. The median growth rate of previous seasons is assumed for the forecast horizon, with admissions calculated based on the growth rate. | Fit jointly across all ICBs. The median growth rate is assumed nationally. Modelled on per capita scale via offset. | Bespoke code & mgcv |

### Scoring rules and scoring analysis data

As our forecasts are probabilistic in nature, we generate entire probability distributions, via quasi-posterior sampling, to provide uncertainty quantification about future values of the target. This set of samples is converted into predictive quantiles; equi-tailed 50% and 90% intervals are reported to users, as well as the median forecast value. However, for evaluation purposes we store equi-tailed intervals with the following nominal coverages: 50%, 90%, as well as predictive medians – the intervals presented to users each week. Proper scoring rules [19] are used to assess probabilistic forecasts in a way which encourages the forecaster to provide an honest assessment of uncertainty about the value to be forecast.

The appropriate scoring rule to be used depends on the nature of the quantity of interest. For our predictions of admissions, we use the per capita weighted interval score (pcWIS). This is the weighted interval score (WIS) applied on a per capita scale, that is, *y* is replaced by *y*′ = *y*/(population size) in the definition of WIS, with quantiles also divided by the population catchment size [20]. Scoring a transformed forecast can be more useful than scoring a raw forecast [21]. This per capita approach allows us to compare scores across different spatial geographies on the same scale, as some have larger population catchments than others e.g. nation versus region. The WIS is a weighted average of interval scores. If a 100(1 − *α*)% prediction interval (*l, u*) is used to estimate *y*′, where 1 − *α* is the nominal coverage, *l* is the lower bound of an interval estimate and *u* is the corresponding upper bound, then the per capita interval score is

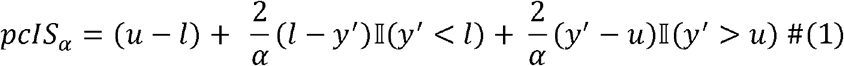

where 𝕀(·) is the indicator function. If we have *K* prediction intervals as estimates of *y*′, as well as a median estimate *m*, then the pcWIS is

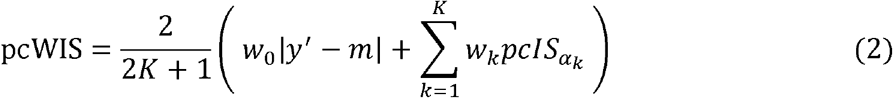

with the usual choice of *w*_*k*_ being 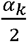, and 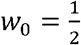 [22]. For pcWIS we evaluate the forecast by averaging across all 14 days of the forecast horizon, as all are shown to forecast users.

The second score we use is for assessing the ordinal component, trend direction, (“increase”, “stable”, “decrease”) of our forecasts using the ranked probability score (RPS) [23]. Let *p*_*j*_ be the forecast probability of category *j* ∈ {1,…,*r*}. Let the observed category be *y* with *o*_*j*_ = 1(*y* = *j*). The ranked probability score is:

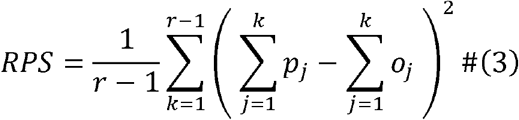

where both pcWIS and RPS are negatively oriented (lower values indicate better performance). For the trend direction scoring we only evaluate the last day of the forecast at 14 days, as the only direction shared with users.

Both pcWIS and RPS are transformed scoring rules with attractive properties which, when taken together, account for different forecast user preferences [21] The pcWIS is an absolute metric, where periods of high values will be penalised more – which is a quality attractive to hospital managers where staff and beds have limited numbers. For example, the growth rate is less useful in this setting than knowing when the quantity of admissions will go above a threshold. The use of a per-capita transformation allows for easier comparison between locations with different population catchments in this absolute scale. The RPS penalises error of the relative prediction, which is a discretised transformation of the epidemic growth rate, which can be more informative to public health officials responsible for interventions.

In all cases we evaluate the predictions against the finalised data set collected after all revisions have been received (14 days following the end of the data forecasted).

### Retrospective simulation experiment

Due to operational constraints, not all the models used in the 2024-25 season were developed or in use at the start of the season, a common challenge for comprehensive evaluation both for in-house modelling efforts and in collaborative hubs where submission is optional. In Figure 1 we see that some models were introduced in the late season, or models were used at different times. Models are sometimes dropped from an ensemble because either the predictions are unrealistic, or an issue which would have been too time consuming to fix before the deadline for forecasts to be shared with users. This means that scores aggregated from predictions over the season do not provide a fair comparison of model performance, as some models are present only in easier periods to forecast. Since only one operational ensemble is used per disease per week, we cannot be certain if we used the “best” possible ensemble, or if individual constituent models were appropriate.

**Figure 1.**
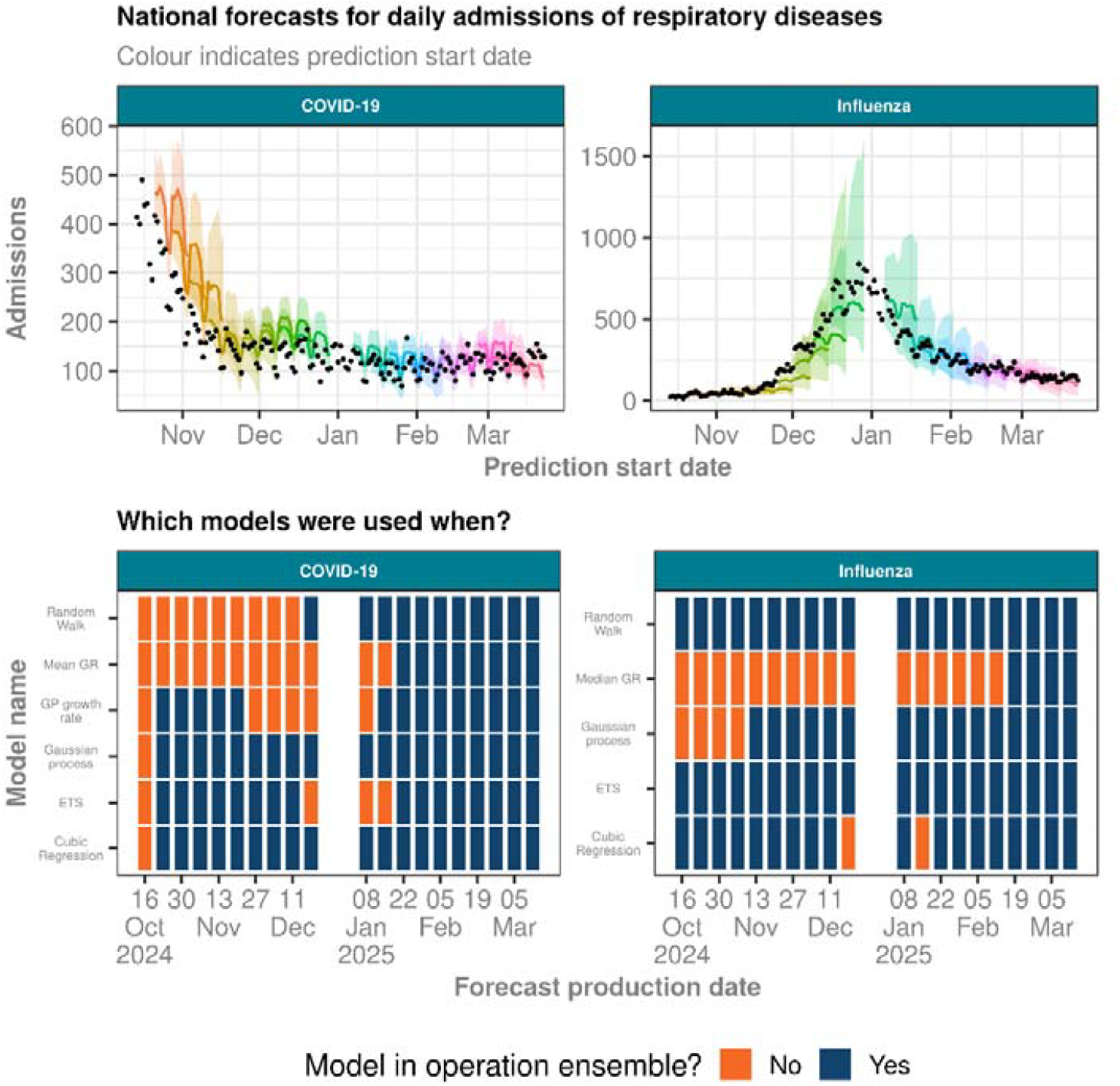
(Top) UKHSA weekly 14-day ahead operational ensemble forecasts for COVID-19 and Influenza, with count of observed admissions shown as black dots. Forecasts are a median prediction (coloured lines) and a 90% prediction interval (colour-matched bands). (Bottom) Graphical representation of which models featured in the operational ensemble over time for each admission target. For operational delivery reasons the Influenza season began a week earlier than for COVID-19.

To help answer these questions we conducted a large simulation experiment. We re-ran each of the models available by the end of the season, producing 14-day forecasts every week from the start of the season until the end of the season. Then, to understand how each model might impact the operational ensemble, we produced all possible sub-ensembles containing 3 models. Computing all possible ensemble combinations would be highly computationally expensive: for a suite of *k* models there are 2^*k*^ − 1 ensembles to be constructed at many spatial geographies. However, the method is general, and any ensemble size is theoretically manageable.

The models and data used to generate these forecasts are based on the final model configurations at the end of the season, and revised data. However, the scale of data revisions is small for this data, with an average 1.3% and 4% change for COVID-19 and Influenza respectively, shown in Supplementary Figure K.

To further reduce computational expense, we used quantile averaging (mean quantile value or mean probability) to produce sub-ensemble forecasts; a common practice for forecasting hubs [5]. In contrast, for operational ensemble forecasts during the season, we constructed an ensemble via unweighted posterior stacking [10, 8]. We then score all models and sub-ensembles via the *scoringutils* package [24].

### Generalised Additive Models (GAMs) and averaging to understand the effect of a model in an ensemble on score

We smoothed the sub-ensemble scores using methods available in the *mgcv* package, and used an averaging method (Equation 1) to estimate the effect of an individual model in a sub-ensemble. This method is used to understand how a model influences the pcWIS as well as RPS. For all scores and diseases, our GAM structure is as follows:

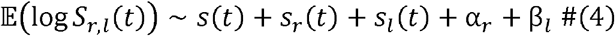

where *S*_*r,l*_ (*t*) is the average score of sub-ensemble *r* for a location level *l* (i. e. nation, region, ICB) over all forecast horizons for a given prediction start date *t, s* (·) is a global smooth for the score over time, *s*_*r*_(·) and *s*_*l*_ (·) are sub-ensemble and location level specific smooths – for each smooth we used penalised thin plate regression splines with knots placed every two weeks – and *α*_*r*_ and *β*_*l*_ are random effects for the sub-ensemble and location level respectively. The structure reflects our forecasting unit: location and time. We use a Gaussian error structure after log-transforming our scores to prevent negative score predictions (pcWIS and RPS are non-negative, real values). We use standard residual-based diagnostics to investigate our GAMs with results in Supplementary Figures M - P of the supplementary information. There are some minor to moderate issues with our GAMs which are a limitation of the analyses relying on the GAMs.

For a given forecasting unit, we define the average effect *p*_*m,l,t*_ of including a model *m* in a sub-ensemble at prediction start date *t* and location level *l* averaged over the forecast horizons as

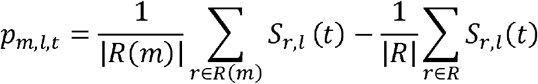

Where *S*_*r,l*_ (*t*) is the mean score of the forecasts across the forecast horizon (days 1 to 14), *R* is the set of all considered sub-ensembles – in our specific case this is all sub-ensembles that can be made of size 3 – and *R*(*m*) is the set of sub-ensembles (of size 3) where *m* is a component model. This is inspired by a functional ANOVA [25] decomposition, and therefore could be extended to understand how combinations of models contribute to sub-ensemble performance. This is, for a fixed location level, the average score from sub-ensembles containing a given model, minus the average score across all sub-ensembles. This is also conceptually similar to the use of Shapley values to understand component model importance [17].

The *gratia* package [26] allows us to generate quasi-posterior samples of *S*_*r,l*_ (*t*) from the models fitted in *mgcv*; we therefore provide median estimates of the effect, as well as 90% credible intervals, by computing each component sum of Equation (5) on a sample-by-sample basis.

### Pareto front

A unique aspect of our forecasting product is the two outputs: a 14-day forecast of target values, as well as the trend direction estimate. This can be viewed as a set of competing objectives, as a single operational ensemble may not strictly optimise both scores. In addition, we have model outputs at different spatial scales. We therefore have 6 competing objectives: pcWIS at national, regional and ICB geographies, and RPS at those same geographies. Optimising performance for a single metric represents an oversimplification of practical forecasting challenges and diverse customer needs.

A Pareto Front (PF) [27] is a tool commonly used when there are multiple competing objectives and no single best solution. Members of a PF can be considered as candidates for a best solution to a multi-objective optimisation problem.

Consider our collection of sub-ensembles *R*, and suppose *r, r*′ ∈ *R* are possible sub-ensembles and *S*_1_ (·), *S*_2_ (·), … are negatively oriented scoring rules applied to the sub-ensembles at a given forecasting unit. We say *r* dominates *r*′ if *S*_*i*_ (*r*) ≤ *S*_*i*_ (*r*′) ∀ *i* and there exists *j* such that *S*_*j*_ (*r*) < *S*_*j*_ (*r*′). We then define the PF as

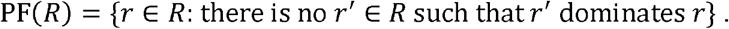

We perform two separate Pareto analyses. First, we perform a simplified Pareto analysis which weights the scores to reduce the dimensionality of the problem: we work with

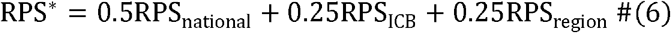

and we weight pcWIS to obtain pcWIS* in the same way. This weighted approach is possible when the scores are on the same scale, which is inherent for RPS, and made possible by using pcWIS rather than WIS. This gives us a two-dimensional front. The weights are chosen to represent a belief that the national forecast is twice as important as the other levels, since this forecast is used by the most senior health officials in the country. However, the other forecasts are not unimportant, since they are used by a wide range of individuals. A “full” Pareto analysis which uses all scores at all spatial geographies is performed with results available in the supplementary materials (Supplementary Table 2).

## Results

### Operational forecasts

Operational ensemble forecasts, and the component models used in each forecast, are presented in Figure 1. Our operational forecasts were able to accurately capture disease dynamics, with 14-day-ahead predictions generated on a weekly cadence. The gap in each set of forecasts is due to a break over the winter holiday period when no forecasts were produced. Supplementary Figure Q shows that the nominal 90% prediction intervals often had close to 90% coverage across diseases and spatial scales. For COVID-19 the forecasts were most closely calibrated after the winter break. For Influenza, the nominal coverage was usually greater than expected, with a drop in coverage in mid-November due to an earlier than expected increase in influenza admissions. Forecasts presented to customers also included regional and ICB breakdowns. Supplementary Figure R shows time series of trend direction estimates. Regional forecasts, and a collection of ICB forecasts are available in Supplementary Figures S and T. Equivalent plots for RSV admissions and geography and age and national Norovirus cases are available in Supplementary Figures A-D.

### Scoring analysis of operational forecasts

For the national operational ensemble forecast, we saw that for COVID-19 the pcWIS generally improved over the season (Figure 2). The slow-changing epidemic dynamics meant accurate forecasts were regularly produced [28]. For Influenza, the pcWIS increases up to the epidemic peak (late December), then decreases again. The number of hospital admissions around the peak of the epidemic was hardest to predict. Our forecast had large uncertainty around the epidemic peak. The predictive intervals were well-calibrated with the nominal coverage: 90% observed coverage statistics were close to the nominal coverage for all diseases and geographies (Supplementary Table 3). However, observed coverage at the 50% level was often higher than expected. The mean pcWIS for the operational ensemble was 4.69 × 10^−7^, which is slightly higher (worse) than the mean for all individual models: 4.61 × 10^−7^. For influenza the mean operational pcWIS was 5.62 × 10^−7^, which is lower than the mean across all individual models: 1.34 × 10^−6^.

**Figure 2.**
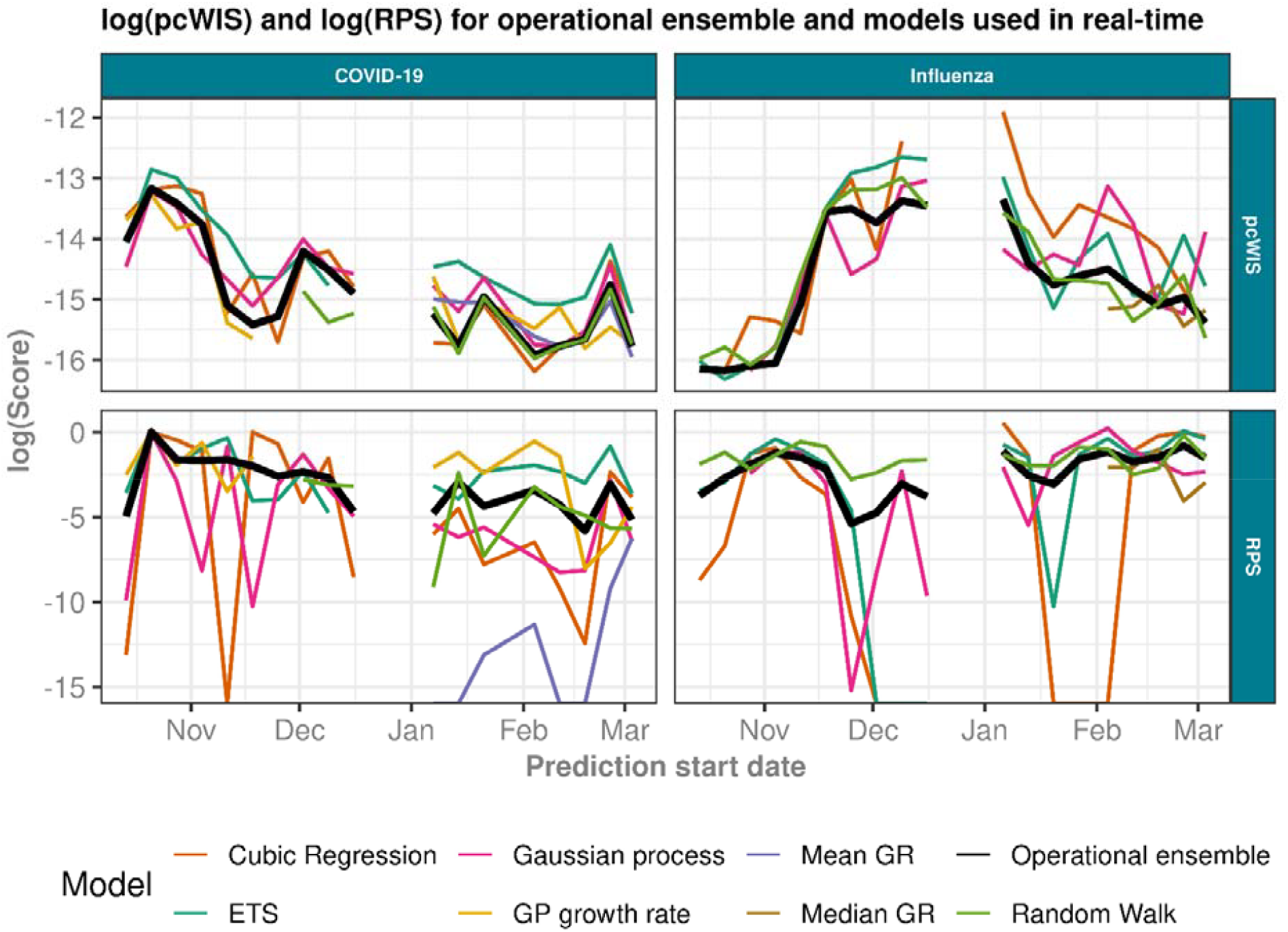
Average scores per prediction start date for COVID-19 and Influenza hospital admissions by model during the 24/25 season. Scores for a given prediction start date are averaged over the 14-day forecast horizons, with lower values indicating better performance. On the top row for each disease is the pcWIS, and on the bottom row the RPS score. The Operational ensemble (the forecast presented to users) is highlighted in black.

In terms of operational ensemble forecast trend direction, RPS was low for COVID-19, often near the theoretical lower bound of 0, with occasional spikes. This indicates the trend direction estimates were typically accurate, but the spikes indicate where trend direction estimation was relatively poor (Figure 2). Again, this is likely due to the simple, stable dynamics of the epidemic. For Influenza, the operational ensemble often performed well, obtaining small RPS values. The performance measure by RPS was consistent near the epidemic peak. As with pcWIS, the RPS values indicate the operational ensembles were, on average, better than individual models. For COVID-19, the mean RPS for the operational ensembles was 0.163, compared to 0.188 for individual models. For Influenza, the mean RPS values for the operational ensemble and individual models are 0.161 and 0.271 respectively. This indicates our ensembles are better at trend estimation than individual models, on average. Scores over time for each model and spatial scale are shown in Supplementary Figure U. To compare the relative skill of forecasts at different population levels we refer the reader to Supplementary Figure V. pcWIS are RPS are both scale-agnostic scoring rules, however, we see that, in general, larger population areas generally correspond to lower (better) score values. This suggests that the larger geographies are usually easier to predict.

### Retrospective simulation analysis of COVID-19 forecasts

Figure 3 shows the effects of individual models on sub-ensemble performance for COVID-19 modelling. For pcWIS, we see that the GP growth rate model was detrimental to sub-ensembles; the pcWIS increased by up to 50% around December, when hospital pressures approached their peak. For RPS, the effects of models on sub-ensemble performance are smoother. We see that, again, GP growth rate is least useful around December-January. However, many other models become less useful as the season progresses. For RPS contribution we see worsening in the performance estimates for ETS, cubic regression and Gaussian process models over the study period. However, random walk and mean growth rate models show improving scores with time. The GP growth rate model is an experimental growth rate model, which can give unusual results when the growth rate is close to 0, which may explain poor performance, particularly around November to December.

**Figure 3.**
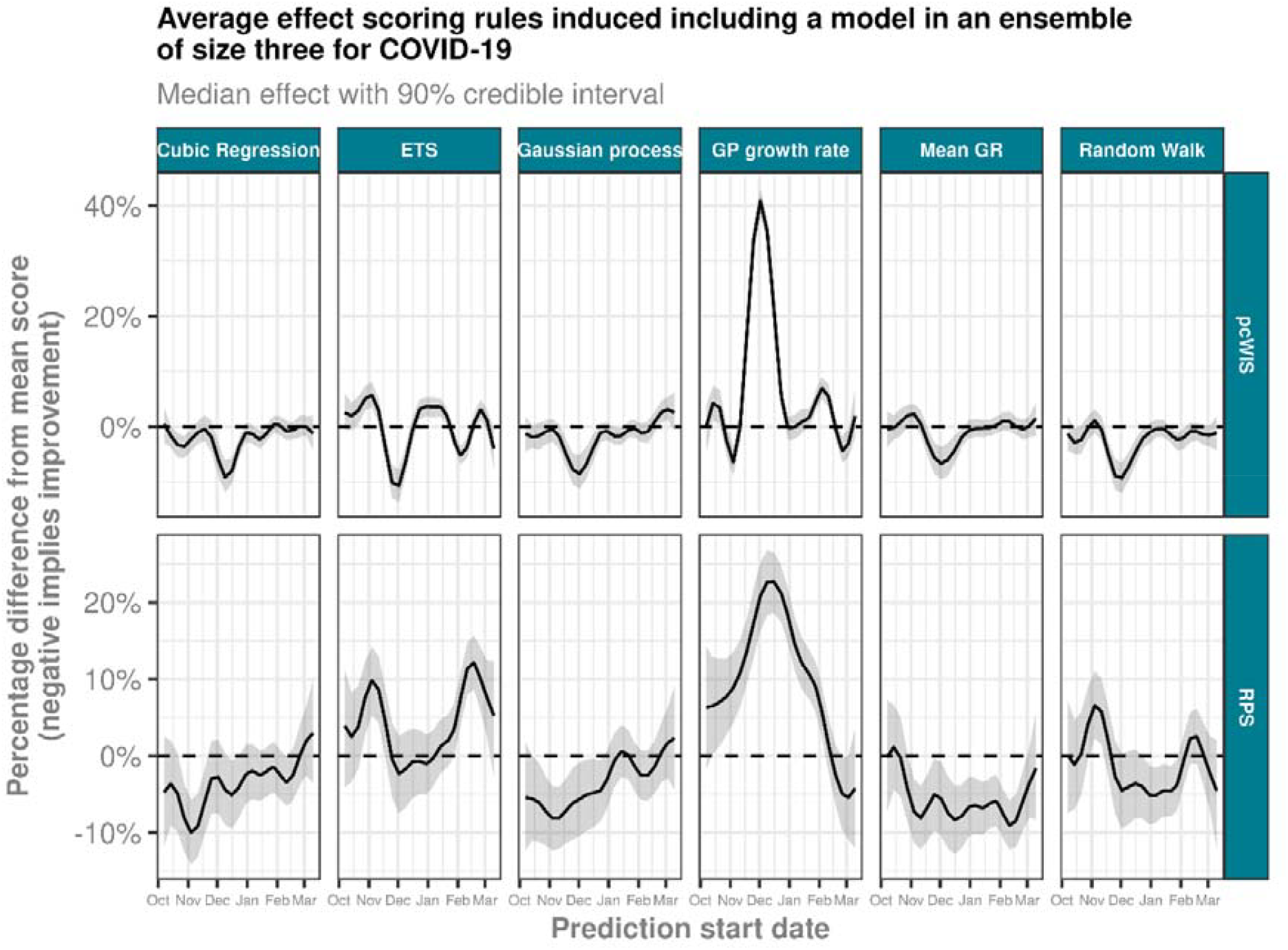
Using data generated from a retrospective evaluation of many sub-ensemble combinations the effect of including a model in an ensemble is estimated using a GAM. Shown are the estimated effects of including a model in an ensemble (expressed as a percentage change in pcWIS or RPS) for COVID-19 forecasts from the overall mean score across all sub-ensembles at that point in time.

However, this did offer better-than-average performance towards the end of the season (see Supplementary Figure W). Comparisons of performance over time for the operational ensembles against individual models at each spatial resolution are given in Supplementary Figure U; the operational ensemble is rarely the best model but typically outperforms most individual models. Season average scores for operational ensembles and individual models are given in Supplementary Table 1 for COVID-19 and Influenza; this shows that the ensemble was on average, provided both better numerical forecasts and trend direction forecasts of COVID-19 and Influenza at all spatial resolutions.

### Retrospective analysis of Influenza forecasts

Figure 4 shows the effects of individual models on sub-ensemble performance for Influenza modelling. We see that the improvement provided by models fluctuates over time. For example, we see that the cubic regression model is worst around the peak (late December – early January): cubic regression models tend to provide approximately exponential dynamics. However, ETS, random walk and Gaussian process models do well around the peak; all of these models have either “flat” predictions, or in the case of the Gaussian process, predictions which revert towards a constant mean. The median growth rate is an experimental model which uses growth rates from previous seasons to provide future forecasts. This provided reasonably good pcWIS contributions. In England, the peak in influenza admissions varies slightly from year to year [29], which may explain relatively poor performance around the peak for the median growth rate model.

**Figure 4.**
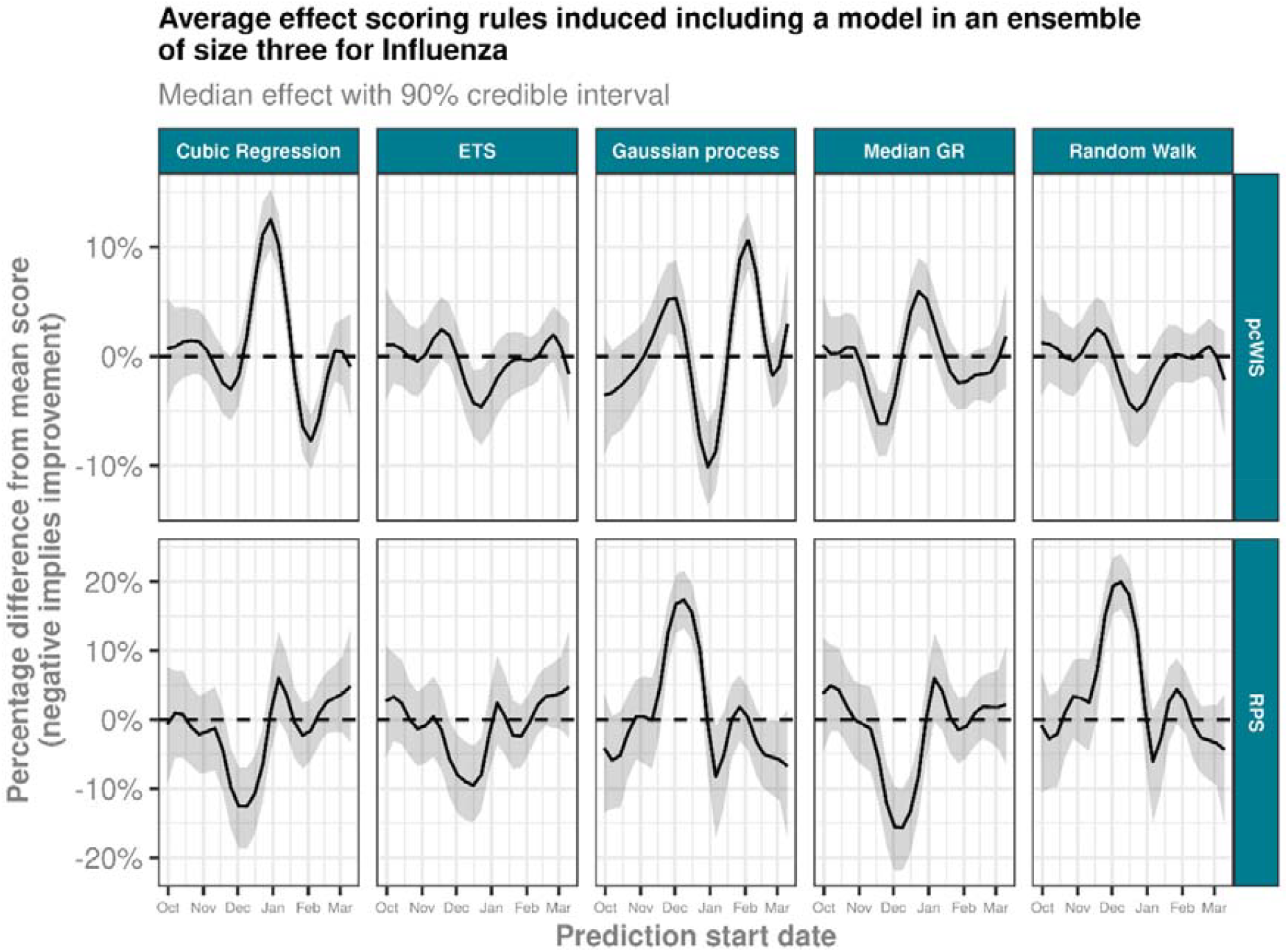
Using data generated from a retrospective evaluation of many sub-ensemble combinations the effect of including a model in an ensemble is estimated using a GAM. Shown are the estimated effects of including a model in an ensemble (expressed as a percentage change in pcWIS or RPS) for forecasts from the overall mean score across all sub-ensembles at that point in time.

### Pareto Analyses

#### Weighted analysis

Since the weighted Pareto analysis is two-dimensional, we can plot the results. Note that in the Pareto analysis, we use an average score over the entire season. This allows us to understand which sub-ensemble combination would have been best over the entire season, on average.

In Figure 5, we see that the PF for COVID-19 is constructed of three sub-ensembles. Each sub-ensemble contains a random walk model. Two contain one of {cubic regression, Gaussian process, mean growth rate}. Since the COVID-19 epidemic was near flat in the 2024-25 season, a random walk component in the ensemble is intuitive. The cubic regression, Gaussian process and mean growth rate models all behave quite differently. We also see a cluster of points far away from the PF. Every model in this sub-ensemble contains a GP growth rate model, suggesting this model is detrimental to average sub-ensemble performance. The values used for the PF analysis are available in Supplementary Figure Y. We see in the Supplementary Figure W of retrospective forecasts that, on occasion, the GP growth rate forecasts exhibited unusual behaviour. However, the difference in score values is typically small between even the best and worst models, when averaging over the season.

**Figure 5.**
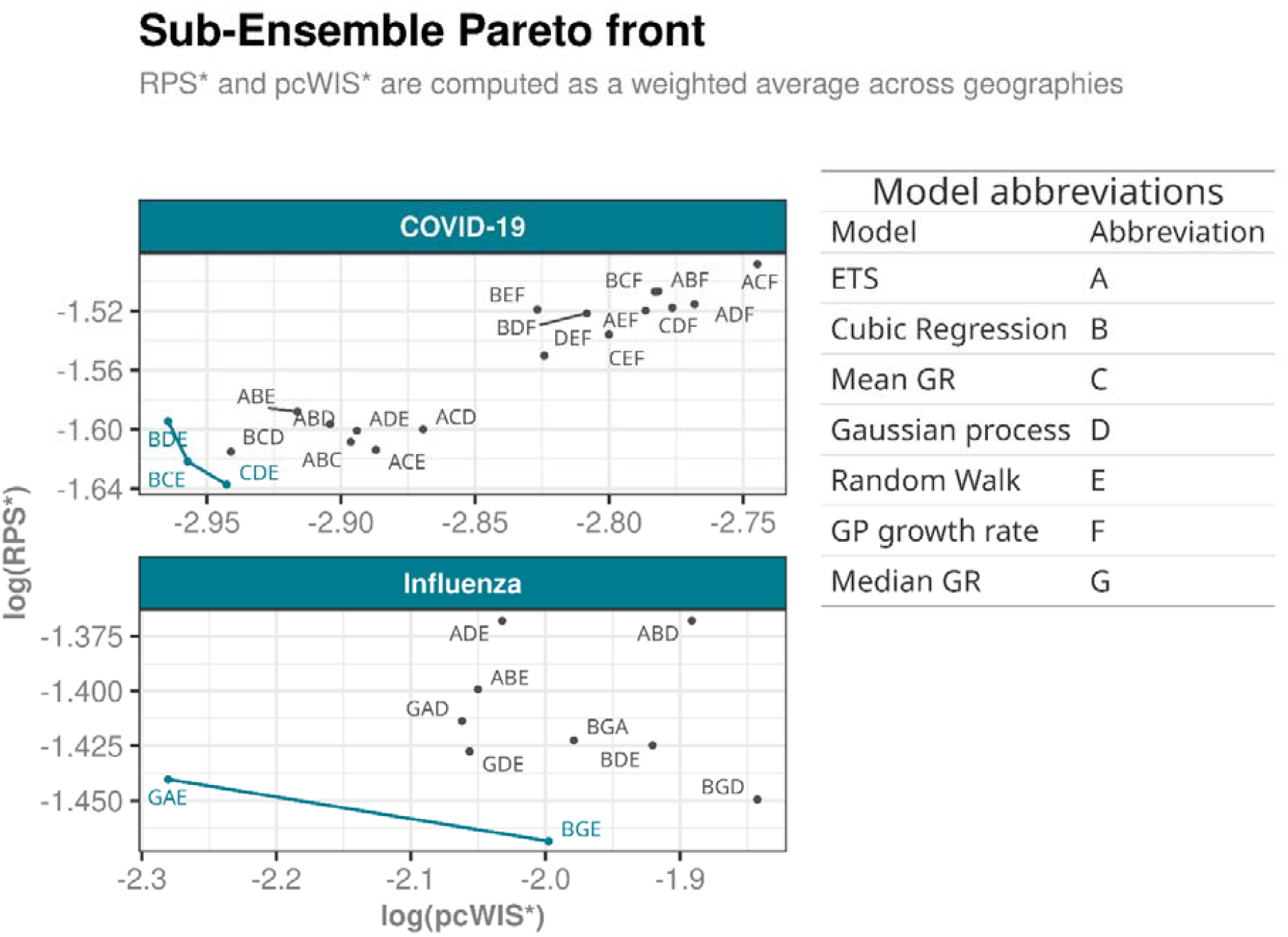
The performance of different sub-ensembles in terms of RPS* & pcWIS*. The sub-ensembles are given abbreviations for clarity to indicate sub-ensemble membership. The scores are generated as the weighted sum of the average value at each geography level (Equation 6). Pareto front for best performing models in pcWIS* and RPS* are shown in blue.

The PF for Influenza under the weighted approach is small. The two members both contain random walk and median growth rate models. Both these models had 90% prediction intervals which almost always contained the true value (Supplementary Figure X), but which were perhaps too wide. The third model in each sub-ensemble (cubic regression or ETS) often had quite narrow prediction intervals. The forecasts for both of these models around the peak could be quite poor but were reasonable in the run up to the peak and then after the peak.

### Comparison of retrospective forecasts to operational forecasts

Our operational ensemble could contain any number of models from one to the number of production models available at that time. Model inclusion in the operational ensemble is a subjective judgement, but we use a model in our weekly operational ensemble if the model predictions are epidemiologically plausible. We see in Figure 6 that, in general, our operational ensemble was of comparable performance to the retrospectively fitted ensembles. We also include a “matched ensemble”. This is an ensemble of the individual retrospective models, where the individual models used to construct the ensemble are the same as the models in the operational ensemble. The matched ensembles typically, but not strictly, outperformed the operational ensembles. This suggests a bias in favour of retrospectively fitted models and sub-ensembles over the operational ensemble as they are fit using revised data and end-of-season hyperparameters. The pcWIS values for the operational ensemble are usually around the average for retrospective sub-ensembles; over the season the average pcWIS values for COVID-19 and Influenza operational ensemble forecasts were 4.69 × 10^−7^ and 5.62 × 10^−7^ respectively. For RPS, the mean RPS scores over the season were 0.188 and 0.271 for COVID-19 and Influenza respectively. Our operational ensembles occasionally gave large RPS values, especially for COVID-19. These spikes in RPS correspond to weeks where the admissions were stable and we forecast a relatively low probability of a stable trend. That being said, if we treat the operational ensemble’s mostly likely trend direction at the national level matched the observed outcome on the majority of occasions: 56% of COVID-19 trend direction forecasts and 63% of Influenza trend direction forecasts matched the observed outcome. Note that the number of correct mostly likely trend direction predictions is not a proper scoring rule, but is an easy to understandable performance metric for a variety of non-technical stakeholders. Table 2 presents results at all spatial resolutions. Our Influenza RPS values for the operational ensemble were often marginally higher than most of the other retrospective sub-ensembles. Minimum mean, overall mean, and maximum mean scores are available in Supplementary Table 4 of the supplementary material.

**Table 2.**
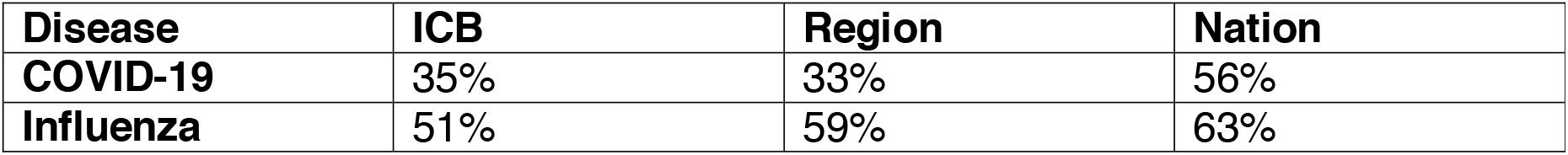
Percentage of occasions the most likely trend direction forecast match the observed outcome by spatial resolution. Influenza forecasts were correct on the majority of occasions. COVID-19 forecasts were not as performant, but were at least as good as random chance (33%) for each geography.

**Figure 6.**
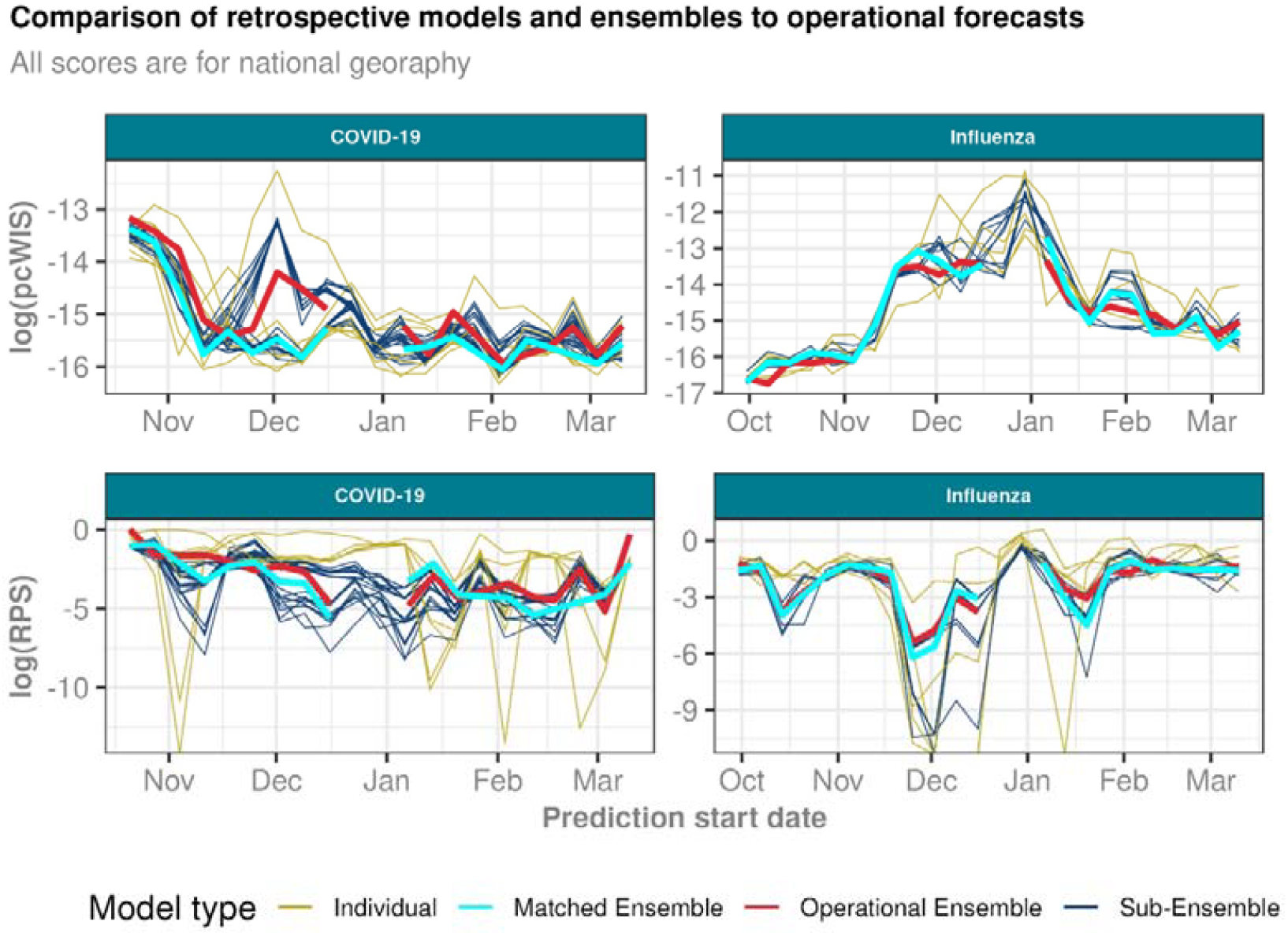
Comparison of how the operational ensemble performs relative to the retrospective 3-model sub-ensembles, the retrospective individual models and the matched ensemble. The scores are presented over time to demonstrate the varying performance at different epidemic phases and at a national geography for pcWIS (top) and RPS (bottom). The break in the red Operational Ensemble and cyan Matched Ensemble lines are due to the forecasting break which occurs over the Christmas and New Year.

## Discussion

Many forecasts used to predict short-term disease dynamics are ensembles. Understanding the component models in such ensembles is important to build better forecasts and divert attention to maintenance and the development of useful models. This paper provides a retrospective evaluation, tooling and methods to understand which models were useful at different epidemic stages and across an entire season for two pathogens.

### Operational Forecasts

Hospital admissions forecasts in the 2024-25 season were broadly accurate. The COVID-19 forecasts were especially accurate; however, the disease dynamics had limited complexity that season. Influenza forecasts were more challenging, especially when the peak was thought to be around the winter break in operations. The operational ensembles are usually better than individual operational models (Figure 2), which is consistent with existing literature [5, 30].

There are numerous challenges to real time modelling including data quality issues and models failing due to bugs found in real-time. Code improvements to our modelling workflow over Summer 2024, most notably using the *targets* package [31], led to an easier-to-maintain collection of modelling pipelines compared to previous years [10]. Another year of experience producing forecasts under pressure led to better real-time decision making; combined with a more mature modelling suite and internal data quality monitoring tools, this resulted in all of our modelling products being delivered on time, or ahead of schedule without failure. We frequently delivered to customers at short notice due to the scheduling of critical system leader meetings.

Notable however, is the Christmas winter holiday period, where there is an agreed pause with users due to changes in surveillance reporting cadences, staff availability and senior officials’ availability. This year, this pause occurred during the peak of the Influenza wave, a period of high-pressure meaning users relied on older forecasts to make decisions. While automation of modelling can improve delivery, capacity of staff trained to assure work is also a key challenge, and forecasts must align to other surveillance data cadences they rely on. The difficulty in this break period is highlighted by the retrospective models poor performance during this time shown in Figure 6, across both pcWIS and RPS.

### Effects of models on sub-ensemble scores

We have proposed a method for discriminating models in ensembles, which we tackled by developing “sub-ensembles”. Similar work includes LASOMO (leave all subsets of models out) and LOMO (leave one model out) methods to understand the importance of models in an ensemble [17]. A novel feature of our approach is to understand how a model may impact multiple different ensembles: the utilisation of GAM smooths can be used to understand the expected improvement in a scoring metric and provide uncertainty about these estimates. This approach gives a detailed understanding of which models are most useful in an ensemble during different epidemic phases, relative to other candidate models. It is possible to estimate component model importance and use this to construct a weighted ensemble, which can have performance benefits [32]; this only shows us which models are useful in relative terms rather than treating models as “good” or “bad”.

When discussing requirements with users of our forecasts there was no clear consensus on what geography or scale (relative or absolute) the forecasts were most important for. This is because the purpose of the forecast varies for the different audience members. For some users the accuracy in terms of absolute values was critical, whereas others found the general direction most important. Users of the forecasts have responsibilities across the national, regional, and local scales. These competing priorities are challenging to directly encode, which motivated our use of a Pareto front analysis. This analysis enables us to highlight the trade-off between different models directly on the front, making the choice between performance for different user types more explicit. The Pareto front gives us a much broader view on the model choice problem than our GAM-based averaging approach on a single evaluation metric alone. An alternative approach to this problem, would be to encode multivariate scoring rules, which should be explored operationally as future work [33].

The geographical weights chosen for the weighted Pareto front analysis are challenging to select and represent our view of the average utility across all users of each spatial level of forecasting. A different opinion of the relative utility of each spatial level may produce a different Pareto front of sub-ensembles. While each user group would prefer higher weighting of the geography most relevant to their work, we assume that the national forecast is used more often by local users than visa-versa to understand overall pressure as well as their own. However, due to aggregation of forecasts and different model structures, not all models perform equally well across spatial scales. Some models assume a national trend in admissions, whereas others assume each ICB is independent which would further drive the possible variation in results due to a different weighting scheme.

We selected a model combination size of 3 models for the sub-ensemble analysis to generate sufficient combinations of models to perform this analysis and have meaningful ensembles. The performance of a given sub-ensemble is a combination of 1) the accuracy & precision of each individual model and 2) the interaction between individual models when ensembled, such as whether model properties such as bias and sharpness cancel. As a result, there should be some similarity between individual models on the Pareto front if the sub-ensemble size were to change. However, the variance of scores across sub-ensembles would decrease as the sub-ensemble size increases. A larger cohort of models would be needed to explore this effect in future work.

Our approach is general and can be applied to any collection of forecasts generated at consistent intervals. The approach is sufficiently flexible to allow users to perform the analysis for different scoring rules, performance metrics or forecasting strata.

We presented a new GAM-based method for understanding how models influence ensemble performance, which can guide decisions about which models can or should be dropped from a modelling suite in favour of new modelling approaches. In particular, as a result of this analysis we have deprecated the COVID-19 Gaussian process growth rate model in favour of alternative approaches. Our Pareto analyses showed us which ensembles were useful when we have multiple scoring criteria which cannot be simultaneously optimised. This is a novel application of the Pareto front to retrospective evaluation of real-time modelling tools.

We also saw that our manually chosen ensemble often performed worse than the retrospectively fitted sub-ensembles. However, observing the published forecasts, our predictions are reasonable. This is reassuring to users of UKHSA forecasting products that the predictions generated are high-quality. However, our operational ensembles often contained more models than the retrospectively fitted sub-ensembles (Figure 1). This shows that the largest possible ensemble, such as those used in forecasting competitions and hubs [34, 5, 35], are not a guaranteed route to improved predictive performance; a recent study advocates for ensembles with components providing complementary information, instead of simply aiming for as large an ensemble as possible [36]. Caution needs to be exercised when opportunistically including models in an ensemble: our ensembles should be diverse and have unique predictive features. This is seen in, for example, our influenza forecasts. It is well-documented in the literature that a diverse ensemble tends to lead to better results [8]. The ETS model tends to have a stable trend and narrow predictive bands, by contrast, the Historic GR model often had a non-stable trend with wide predictive intervals. Other influenza models had different behaviours to these models (Supplementary Figure T).

This research has shown that the complex biological systems of infections over time are difficult to forecast consistently well, with varying model performance over time – shown by the GAM based contribution analysis. Furthermore, we showed that different aspects of the system – absolute scores vs relative change are not equally easy to forecast under each model, with different combinations of models having different skill, as demonstrated by the Pareto Front analysis. We showed that models which depend on growth rate assumptions tend to add value in incline and decline phases, whereas models that assume time series trends or flat forecasts can improve performance when epidemic waves are more stochastically variable. Having the tools available to inspect different aspects of ensemble performance allows public health forecasters to evaluate more rigorously, and therefore draw better conclusions to plan future model combinations.

### Limitations

Limitations of our approaches are as follows. We have only used pure-statistical approaches in these ensembles. The ensemble could be improved with the addition of semi-mechanistic approaches which include parameters such as susceptible population sizes and infection hospitalisation rates. Semi-mechanistic approaches are distinct addition to our future ensembles [37, 38]. Bespoke solutions can be complex to develop, but packages such as *EpiNow2* provide multiple modelling options [39]. As with any scoring-based approaches, this work is retrospective, thus cannot be used in real time. However, scoring can be used mid-season to assess relative performance of models and contribution to model performance. Although there is evidence in the literature to suggest 4 or more models construct a robust ensemble [40], we chose sub-ensembles of size 3 due to the limited number of models available for combining.

The GAMs fit to scores for Figures 3 and 4 to understand how individual models contribute to sub-ensemble performance, like all models, have their limitations. Diagnostic plots for the GAMs are given in Supplementary Figures M-P. Future work should explore strength and limitations of the used of statistical model based ensemble analysis.

The (relatively) poor performance of our operational ensembles compared to retrospective sub-ensembles can be attributed to a few factors. Firstly, not all models were available throughout the season, with some being introduced very late in the season when we had more time to experiment with methods. The matched ensemble shows that even using the same models as in real-time, the retrospective individual models outperformed the operational ensemble. The 2024-25 season posed new challenges for the team as we employed new technologies to increase operational efficiency but simultaneously were often under pressure to deliver forecasts ahead of regular cadence to meet decision makers’ needs. This meant the team had less time to choose and experiment with different operational ensembles, as we had to provide forecasts to customers promptly. We were able to apply expert judgement to our forecasts and tune hyperparameters accordingly, and to include/drop models from our operational ensembles; for COVID-19 this led to comparable performance when comparing the matched ensemble to individual models and sub-ensembles. For Influenza, the matched ensemble often outperformed individual models and sub-ensembles (Figure 6). Furthermore, data is revised in real-time which can degrade model performance operationally when compared to models forecast using retrospectively available data.

Our future work will be to diversify our ensembles by including infection dynamic mechanism, as well as using models which use more long-term historic data to use information such as seasonality to improve modelling. This will allow us to have a rich ensemble at the start of the season and provides a suite of modelling tools for use in the event of an incident or unexpected pandemic. We will expand RSV and norovirus operational ensembles to gain the benefits which ensembles can bring to performance. This will allow us to perform similar analyses for those diseases.

We have already removed models from our suite based on this analysis and could use this analysis to further improve future operational ensembles. Our operational ensembles are currently unweighted, thus one idea is to construct ensemble weights for a future season or dynamically within season [41, 42]. For example, the *p*_*m,l,t*_ values could be used to calculate time-varying ensemble weights [8]; one challenge will be understanding how to use *p*_*m,l,t*_ values for different scoring rules, forecasting outputs, or spatial resolutions to construct a single set of weights. As the size of our ensemble grows, we can use our Pareto analyses as a first-pass for model evaluation: models which do not feature in a Pareto front should be considered for deprecation.

## Acknowledgements

We thank UKHSA data operations colleagues for their work in the access, processing and maintenance of the data used in this paper. We would also like to thank Maximillian Ayling, Gregory Barnsley, Phoebe Asplin, and Ian McFarlane of the Infectious Disease Modelling team for contributing to the operational delivery of forecasts during the Winter 2024-25 season.

## Author contributions

Jack Kennedy: Conceptualisation; Methodology; Software; Formal analysis; Writing – Original Draft; Writing - Review & Editing; Visualization. William Ferguson: Software; Writing - Review & Editing. Owen Jones: Software; Writing - Review & Editing; Data curation. Steven Riley: Writing – Review & Editing. Thomas Ward: Writing – Review & Editing. Maria Tang: Software; Writing - Review & Editing. Jonathon Mellor: Conceptualisation; Methodology; Software; Validation; Writing - Original Draft; Writing - Review & Editing; Supervision.

## Data availability

An application for data access can be made to the UK Health Security Agency. UKHSA operates a robust governance process for applying to access protected data that considers:

- the benefits and risks of how the data will be used
- compliance with policy, regulatory and ethical obligations
- data minimisation
- how the confidentiality, integrity, and availability will be maintained
- retention, archival, and disposal requirements
- best practice for protecting data, including the application of ‘privacy by design and by default’, emerging privacy conserving technologies and contractual controls.

Access to protected data is always strictly controlled using legally binding data sharing contracts. UKHSA welcomes data applications from organisations looking to use protected data for public health purposes. To request an application pack or discuss a request for UKHSA data you would like to submit, contact.

The modelling code for this project is available at https://github.com/jcken95/sub-ensemble-evaluation.

## Supporting information captions

*Figure A. National norovirus case forecasts by prediction start date. Forecasts are for a 14-day horizon, with nowcasts not visualised. A single model was used rather than an ensemble. Norovirus cases increase over the season. Like other pathogens, there is a two-week break in forecasting over the Christmas period*.

*Figure B. RSV national operational forecasts (top) and model used (bottom). The first 6 RSV forecasts were not ensembles, they were single model forecasts (first a thin-plate spline, then a Gaussian process). The thin plate spline model was deprecated entirely early in the season due to beliefs about forecast credibility*.

*Figure C. RSV ensemble forecasts by NHS region. Black dots are daily admissions of RSV hospital admissions per day. Each region has similar disease dynamics: a peak in RSV admissions in early December, with a gradual decline in admissions. Incidence in each region peaks between 50 and 250 admissions*.

*Figure D. RSV operational ensemble admissions forecasts by age group (national geography). Daily admissions are shown as black points. Incidence varies greatly with each age group. The [0,2) group peaks in early December at just below 500 daily admissions, whereas the [65,75) age group peaks at around 70 daily admissions in late December. In general, younger age groups peak earlier than the older age groups*.

*Figure E. log(pcWIS) per prediction start date for norovirus cases single model forecasts and RSV operational ensemble admissions forecasts at national geography. Model performance is variable for Norovirus. RSV admissions predictions are generally better after the Christmas break than before the Christmas break. The RSV epidemic is generally decreasing after the Christmas period*.

*Figure F. log(pcWIS) per prediction start date and NHS Region for RSV operational ensemble admissions forecasts. The performance of the ensemble is similar across regions, with each region showing increasing performance as we progress through the season. This increase in performance could be attributed to (i) reduced incidence and (ii) improvements in the ensemble over the season*.

*Figure G. log(pcWIS) by age group (national geography) and prediction start date for RSV operational ensemble forecasts. Performance is generally worse for the very young, or very old age groups who typically have higher incidence. Within age groups, forecast performance tends to increase slightly as we progress through the season. This could be attributed to (i) reduced case counts (ii) development of the ensemble in real time*

*Figure H. log(RPS) by prediction start date for national single model Norovirus cases and national RSV operational ensemble forecasts. Norovirus scoring starts later in the season because (i) norovirus modelling started later in the season and (ii) trend direction estimation for norovirus was developed later in the season*.

*Figure I. log(RPS) by prediction start date and NHS Region for RSV operational ensemble admissions forecasts. Trend direction performance was similar across the season, with some early season estimates offering mixed results*.

*Figure J. log(RPS) by prediction start date and age group (national geography) for RSV operational ensemble admissions forecasts. Trend direction estimation performance was similar across the season, with some correct, and highly confident, forecasts early in the season in the two youngest age groups*.

*Figure K: (Top) A time series of admissions over time at the national level for COVID-19 and Influenza. The red line indicates the data UKHSA received initially, and in blue the final revised version of the data. (Bottom) The percentage change from the initially received admission count to the final counts over time for COVID-19 and Influenza. As national data is shown for simplicity, some local variation may be hidden due to cancelling directions. There are larger percentage changes for influenza at the start and end of the season where counts are low. The revisions for Influenza are largest near the winter holiday period, where operational forecasts were not reported*.

*Figure L: Map illustrating NHS regions and ICBs in England, and their approximate population sizes. The smallest geographies are coloured by their population sizes. The NHS regions and noted by black bold lines. London, due to its relatively small geographical area, is shown in a cut-out in the top right of the figure. Data processed and figure generated using R, using spatial boundaries and population catchment data. Source: Office for National Statistics licensed under the Open Government Licence v*.*3*.*0. Contains OS data © Crown copyright and database right 2026. National boundaries: https://geoportal.statistics.gov.uk/datasets/ons::countries-december-2024-boundaries-uk-buc-2/about; regional boundaries: https://geoportal.statistics.gov.uk/datasets/ons::nhs-england-regions-january-2024-boundaries-en-bfc/about; ICB boundaries: https://geoportal.statistics.gov.uk/datasets/ons::integrated-care-boards-april-2023-boundaries-en-bsc/about*

*Figure M: Diagnostic plots for COVID-19 pcWIS GAM. QQ plot shows moderate deviation from Normality. Histogram shows approximately symmetric residuals and suggests heavy-tailed residuals. Observed vs fitted values shows an approximately linear relationship, with a small cluster of outliers. The Residuals vs linear predictor shows three main groups (one per location level) and a small cluster of potential outliers. Overall, there is moderate deviation from Normality and homoscedasticity of residuals which may affect inferential statements, but predicted values are reasonably close to observed values. This is expected to have only a minor impact on pcWIS analysis*.

*Figure N: Diagnostic plots for COVID-19 RPS GAM. QQ plot shows moderate deviation from Normality. Histogram shows approximately symmetric residuals, there is mild assymetry and heavy-tailed residuals. Observed vs fitted values shows an approximately linear relationship, there is some mild curvature and moderate scatter. The Residuals vs linear predictor shows a decrease in residual variability as the value of the linear predictor increases, but no other obvious patterns. Overall, there is moderate deviation from Normality and homoscedasticity of residuals which may affect inferential and predictive statements. This could have a moderate effect on inferences of the COVID-19 RPS analysis*.

*Figure O: Diagnostic plots for Influenza pcWIS GAM. QQ plot shows moderate deviation from Normality. Histogram shows mildly asymmetric residuals. Observed vs fitted values shows an approximately linear relationship, with a small group of potential outliers when the fitted values are large. The Residuals vs linear predictor some evidence of residual variance increasing with the linear predictor. Overall, there is mild deviation from Normality and homoscedasticity of residuals which may affect inferential statements, but predicted values are reasonably close to observed values. This is expected to have only a minor impact on Influenza pcWIS analysis*.

*Figure P: Diagnostic plots for Influenza pcWIS RPS. QQ plot shows moderate deviation from Normality with one point having a very large, negative residual (approximate value, −6*.*5). Histogram shows mildly asymmetric residuals. Observed vs fitted values shows an approximately linear relationshipbut with variable scatter. The Residuals vs linear predictor some evidence of residual variance decreasing with the linear predictor. Overall, there is mild deviation from Normality and homoscedasticity of residuals which may affect inferential statements, but predicted values are reasonably close to observed values. The large potential outlier was for the gam_cr_gam_gp_ets sub-ensemble. This had a perfect RPS score of 0 which causes problems with a log transform. A small offset* (10^−6^) was added to all RPS values in the Influenza analysis to avoid *log* (0). However, the GAM still struggled to adequately account for this value. Overall, this is expected to have a moderate impact on Influenza pcWIS analysis.

*Figure Q: Comparison of observed and nominal coverage statistics for forecasts by disease and spatial granularity, averaged over the 14 day prediction horizon. No bands are present for the national forecasts as there is only one national forecast per prediction start date-disease combination. The orange bands for Region and ICB represent the min and max value present across the locations. The blue, horizontal line represents the nominal coverage (90%)*.

*Figure R: (Top) Trend direction probability estimates for national COVID-19 and Influenza admissions forecasts of the operational ensemble at national geography. (Bottom) Observed epidemic trend directions at national geography*.

*Figure S. Regional operational ensemble forecast for COVID-19 and Influenza. In these cases, the shapes of the admissions curves are similar to the corresponding national forecasts, but naturally, the number of admissions is lower at finer spatial resolutions*.

*Figure T: Example ICB forecasts for daily COVID-19 and Influenza hospital admissions. ICB names are redacted for data governance reasons. Like regional forecasts, ICB forecasts tend to follow a similar epidemic curve to the corresponding national forecast, but the scale of the epidemic is much smaller*.

*Figure U. log(pcWIS) and log(RPS) per disease-location level combination. Note for some individual models, the trend direction estiamte was near perfect, so the log scores diverge to* −∞ *and are therefore not visible on the plot*.

*Figure V. (Top) Bubble chart showing log(RPS) vs log(pcWIS) for different forecasting locations, point size represents the population of the location for forecasting. (Bottom) Boxplot showing population sizes per location level. In general, scores are lower (better) for locations with a larger population*.

*Figure W. Individual retrospective model predictions for COVID-19 hospital admission at national geography. The national wave during the study period included a decline then stable period with limited variation. Most models capture these dynamics reasonably well, but the gr_gp model is highly erratic*.

*Figure X. individual retrospective model predictions for Influenza hospital admissions at national geography. The epidemic wave showed a sharp rise in admissions followed by a more gradual decline. Most models capture the general epidemic shape, but with high uncertainty. The gam_gp model appears to be very poor in many cases*.

*Figure Y. Values used in the Pareto front calculations (Figure 5)*.

*Supplementary Table 1. Mean scores for models used in winter operations and the matched ensemble. Smaller values indicate better forecasts. In all cases, the average ensemble score for each location level-disease-score group was less than the average operational individual models score. The matched ensemble was usually the lowest scoring in each location level-disease-score group; the operational ensemble was the best for Influenza pcWIS at region and ICB level*.

*Supplementary Table 2. Members of Pareto Front for full Pareto analysis (not using weighted averages) by disease*.

*Supplementary Table 3. Comparison of nominal coverage of prediction intervals to observed coverage by disease and spatial granularity for operational ensembles. Statistics are an average for the entire season and where relevant, averaged across locations*.

*Supplementary Table 4. Minimum, overall mean, and maximum values of retrospective ensemble scores averaged across the season for national predictions; with operational ensemble average for comparison*.

*Supplementary Table 5. Mean scores for models used in winter operations. Smaller values indicate better forecasts. In all cases*.

*Supplementary Section B: A specification of the novel models added in the 2024-2025 season for COVID-19 and Influenza forecasts. Hyperparameters are provided within the configuration files in https://github.com/jcken95/sub-ensemble-evaluation.*

