## supplementary information for "Evaluation of short-term multi-target respiratory forecasts over winter 2024-25 in England using sub-ensemble contribution analyses"

Supplementary information to the paper: “*Evaluation of short-term multi-target respiratory forecasts over winter 2024-25 in England using sub-ensemble contribution analyses*”

Jack Kennedy^1^, William Ferguson^1^, Owen Jones^1^, Steven Riley^1,2^, Thomas Ward^1^, Maria L. Tang^1^, Jonathon Mellor^1^

1. UK Health Security Agency, London, UK
2. School of Public Health, Imperial College London, London, United Kingdom

### Supplementary Section A

### RSV admissions and Norovirus cases analysis

RSV admissions are forecast at the regional level, whilst also accounting for the age group of the admitted patient. Forecasts are then aggregated to one of three levels for reporting (and evaluation) purposes:

- National forecast (all ages)
- National forecast (by age group)
- Regional forecast (all ages)

The regions are the same NHS regions as for COVID-19 and Influenza forecasts. Age groups are [0, 2), [2, 5), [5, 18), [18, 65), [65, 76), and [75, 120) years old. These groups were selected with the help of epidemiologists with expertise in RSV and winter pathogens.

Norovirus cases are a joint nowcast and forecast model see (cite) for details. In the 24/25 season, norovirus modelling was done only at the national scale and the forecast was a single model. By the 25/26 season this was developed into a regional ensemble model. We also note that storage of norovirus results started later in the season compared to other pathogens as it was a new forecasting target for the 24/25 season.

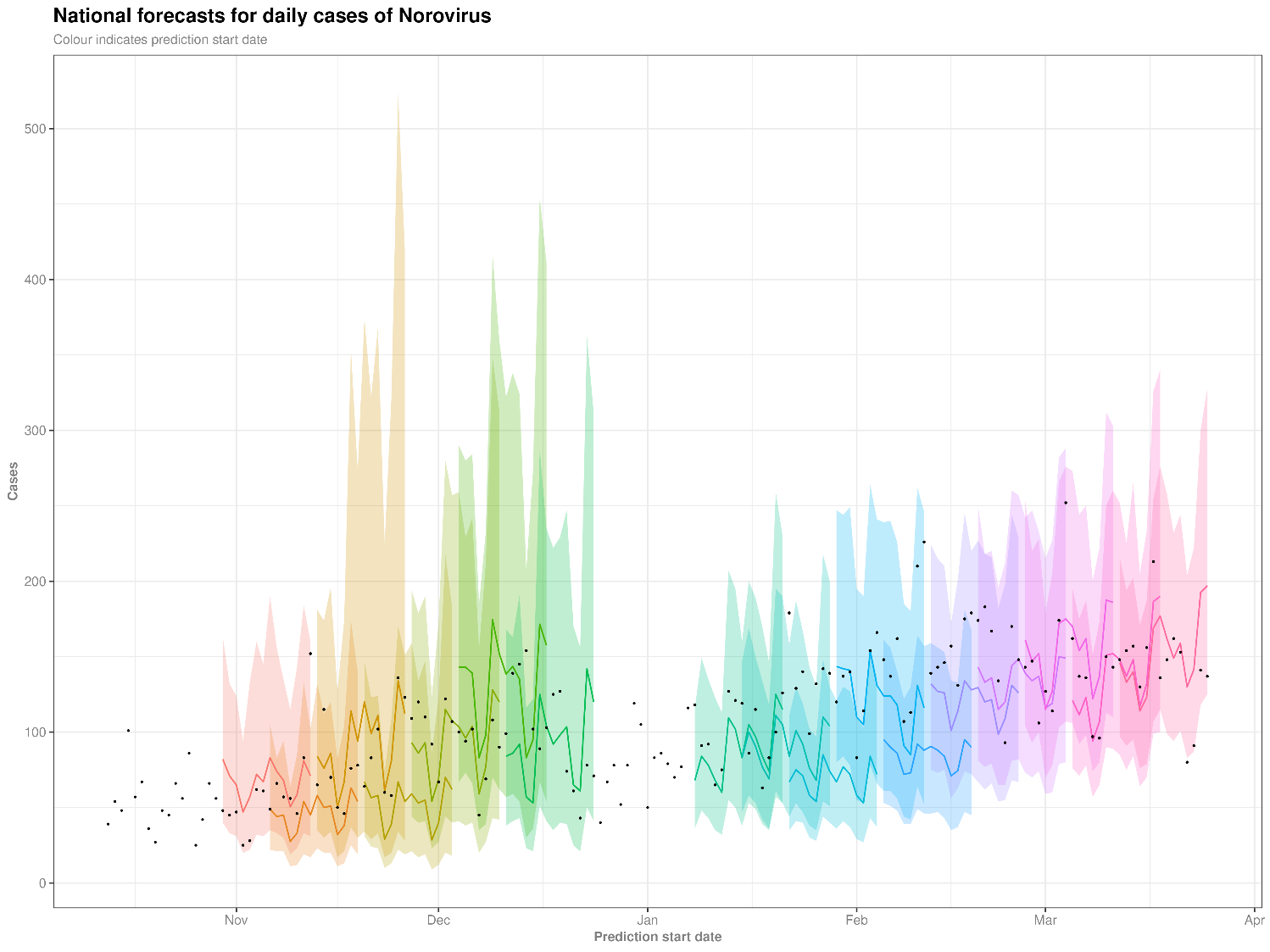

Figure A. National norovirus case forecasts by prediction start date. Forecasts are for a 14-day horizon, with nowcasts not visualised. A single model was used rather than an ensemble. Norovirus cases increase over the season. Like other pathogens, there is a two-week break in forecasting over the Christmas period.

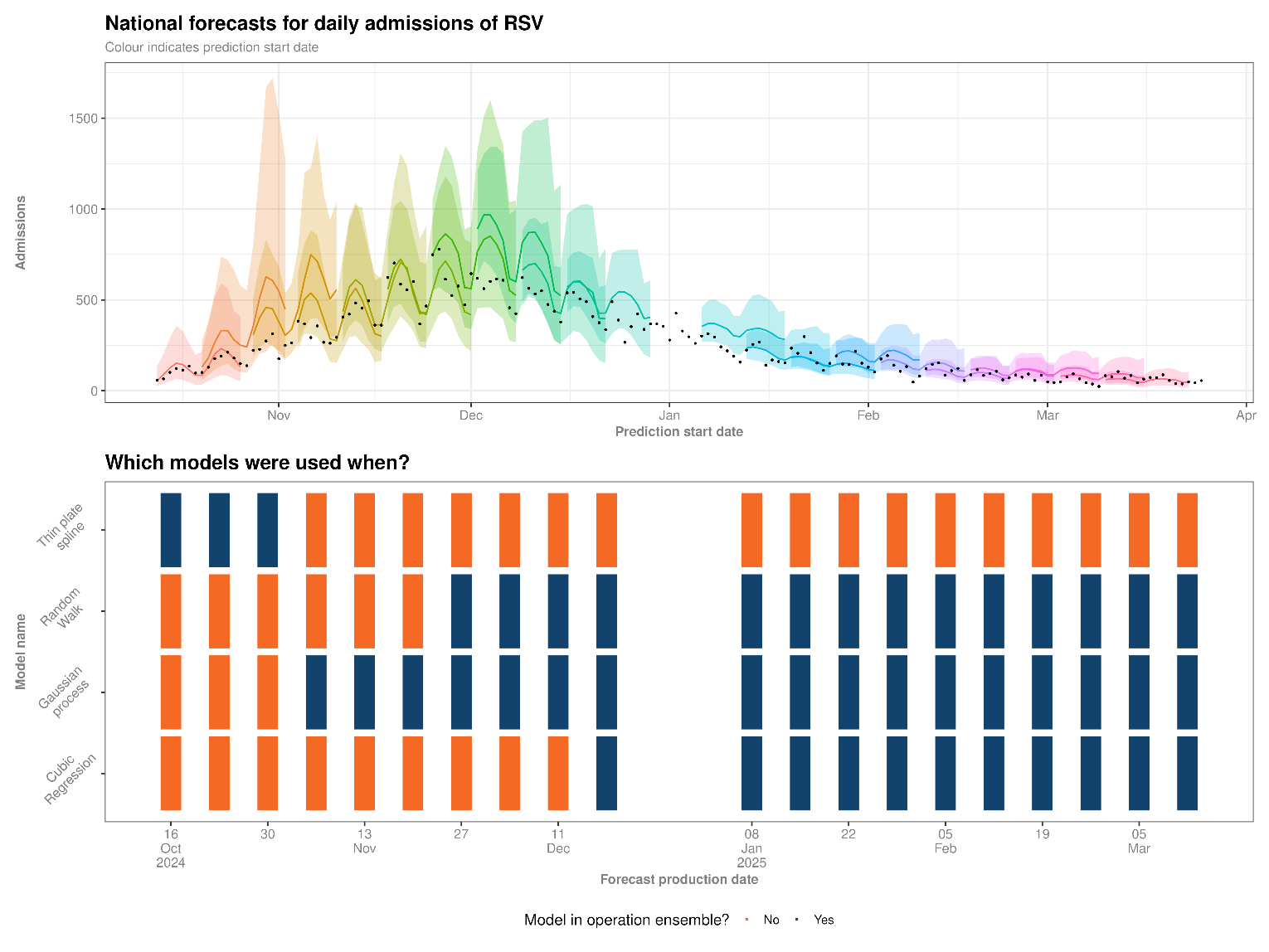

Figure B. RSV national operational forecasts (top) and model used (bottom). The first 6 RSV forecasts were not ensembles, they were single model forecasts (first a thin-plate spline, then a Gaussian process). The thin plate spline model was deprecated entirely early in the season due to beliefs about forecast credibility.

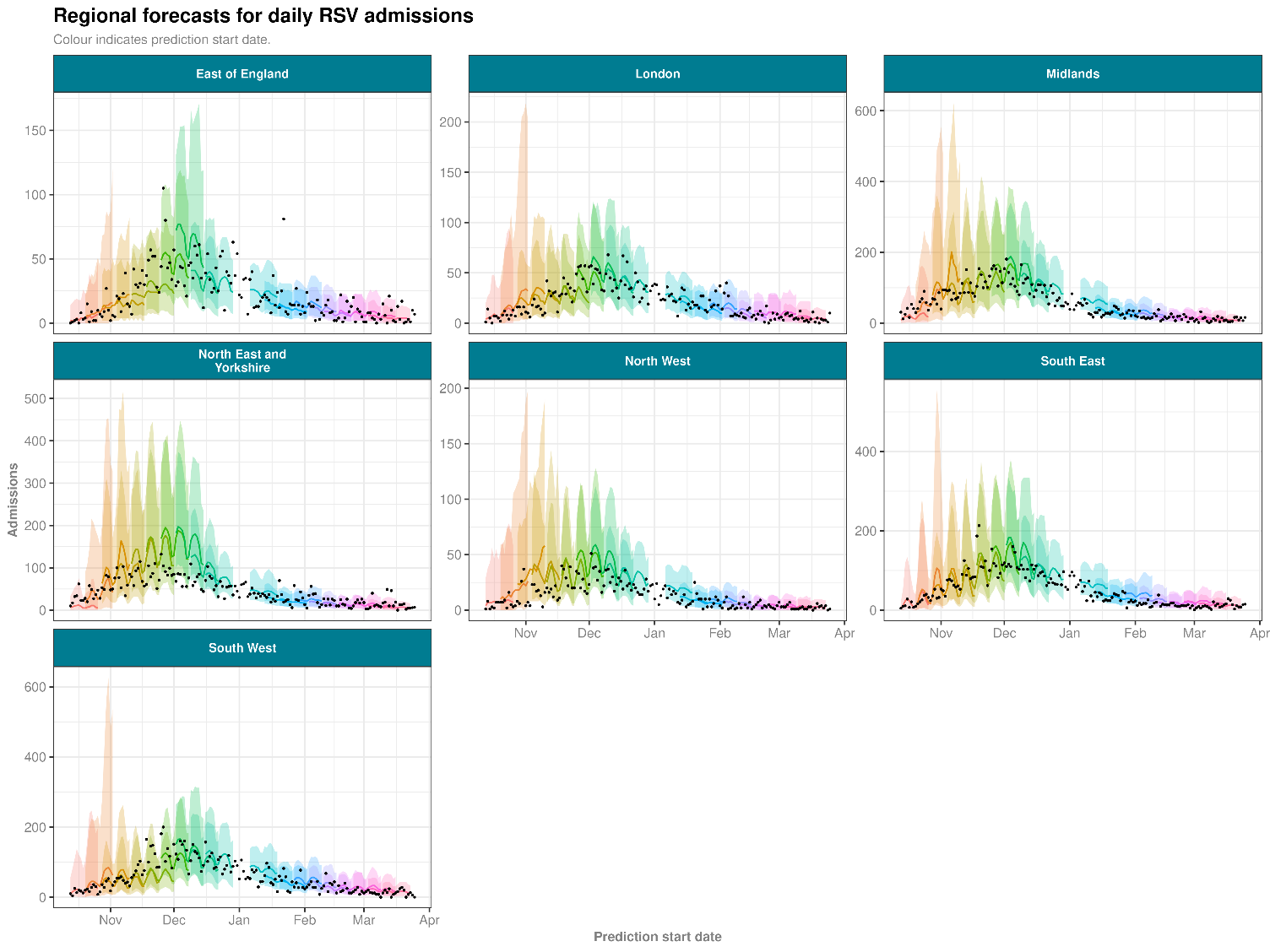

Figure C. RSV ensemble forecasts by NHS region. Black dots are daily admissions of RSV hospital admissions per day. Each region has similar disease dynamics: a peak in RSV admissions in early December, with a gradual decline in admissions. Incidence in each region peaks between 50 and 250 admissions.

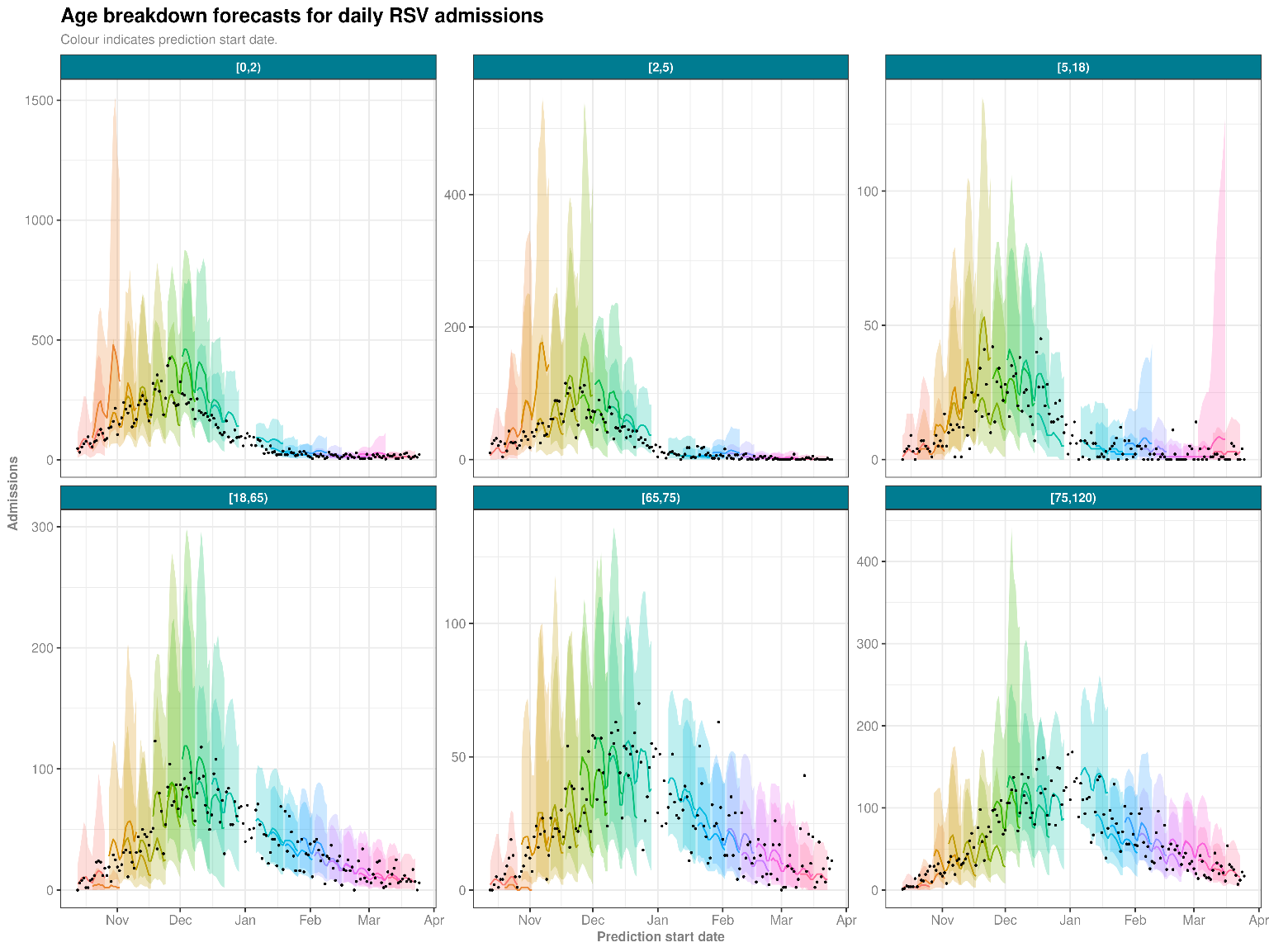

Figure D. RSV operational ensemble admissions forecasts by age group (national geography). Daily admissions are shown as black points. Incidence varies greatly with each age group. The [0,2) group peaks in early December at just below 500 daily admissions, whereas the [65,75) age group peaks at around 70 daily admissions in late December. In general, younger age groups peak earlier than the older age groups.

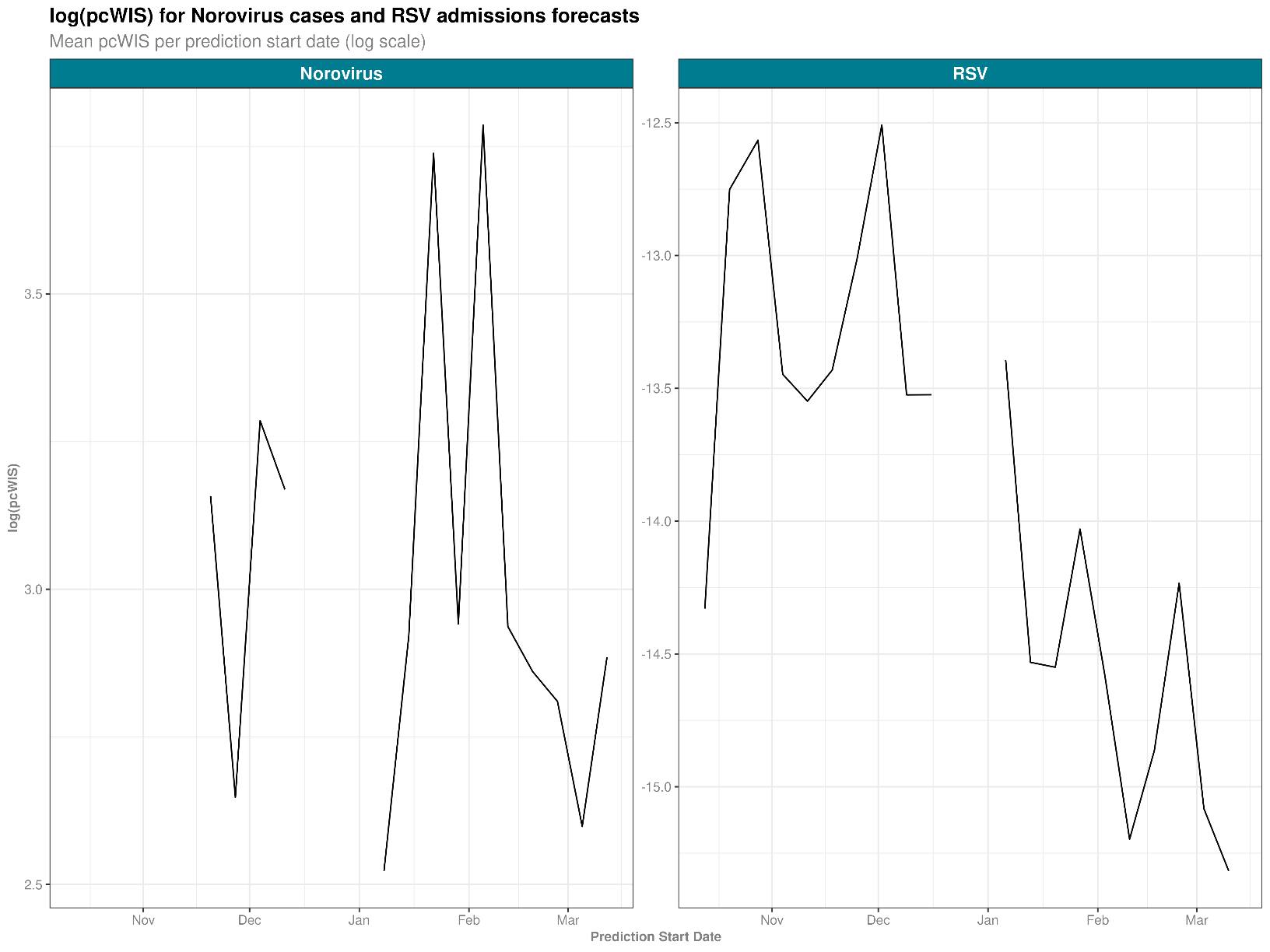

Figure E. log(pcWIS) per prediction start date for norovirus cases single model forecasts and RSV operational ensemble admissions forecasts at national geography.

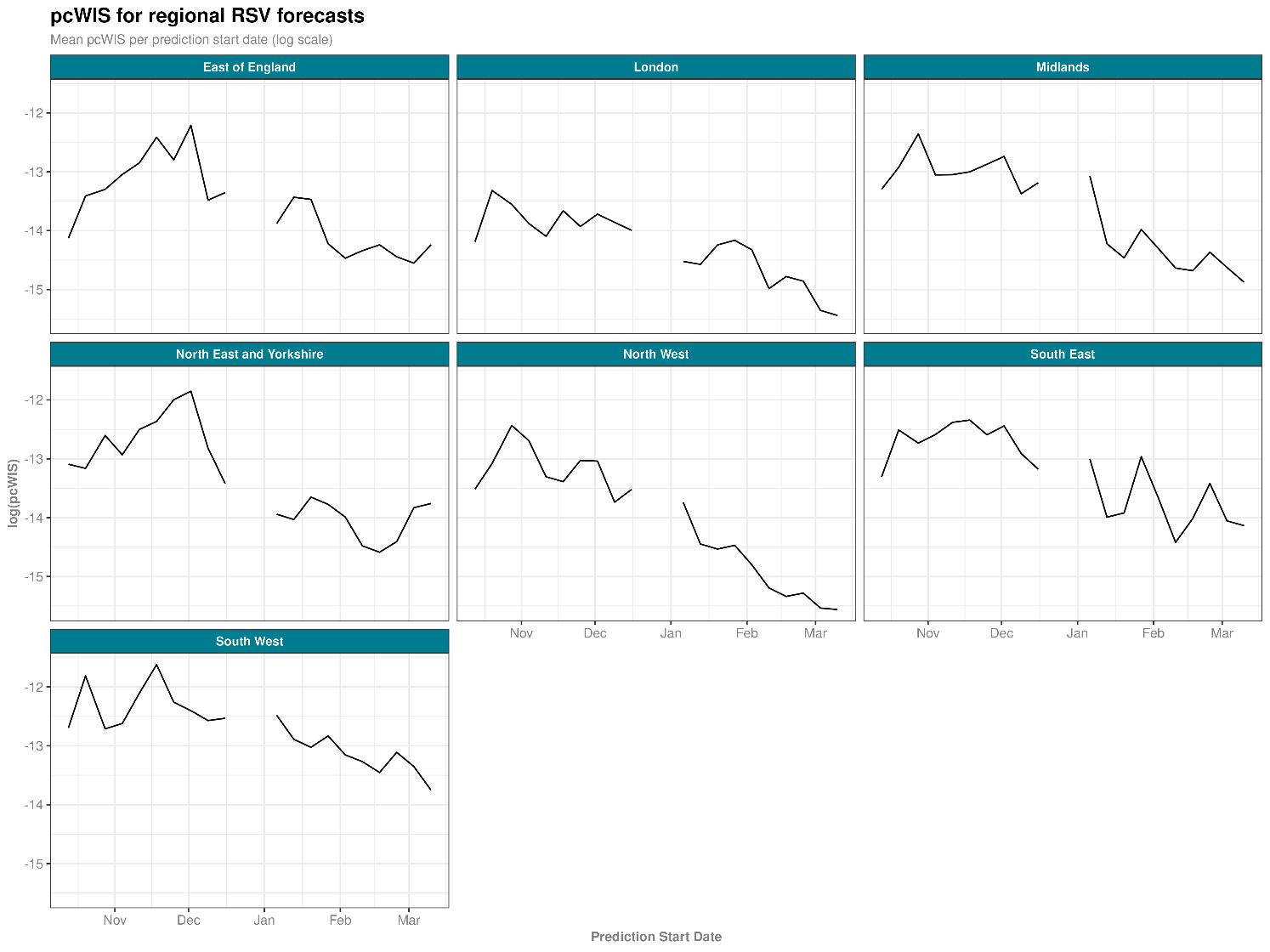

Figure F. log(pcWIS) per prediction start date and NHS Region for RSV operational ensemble admissions forecasts. The performance of the ensemble is similar across regions, with each region showing increasing performance as we progress through the season. This increase in performance could be attributed to (i) reduced incidence and (ii) improvements in the ensemble over the season.

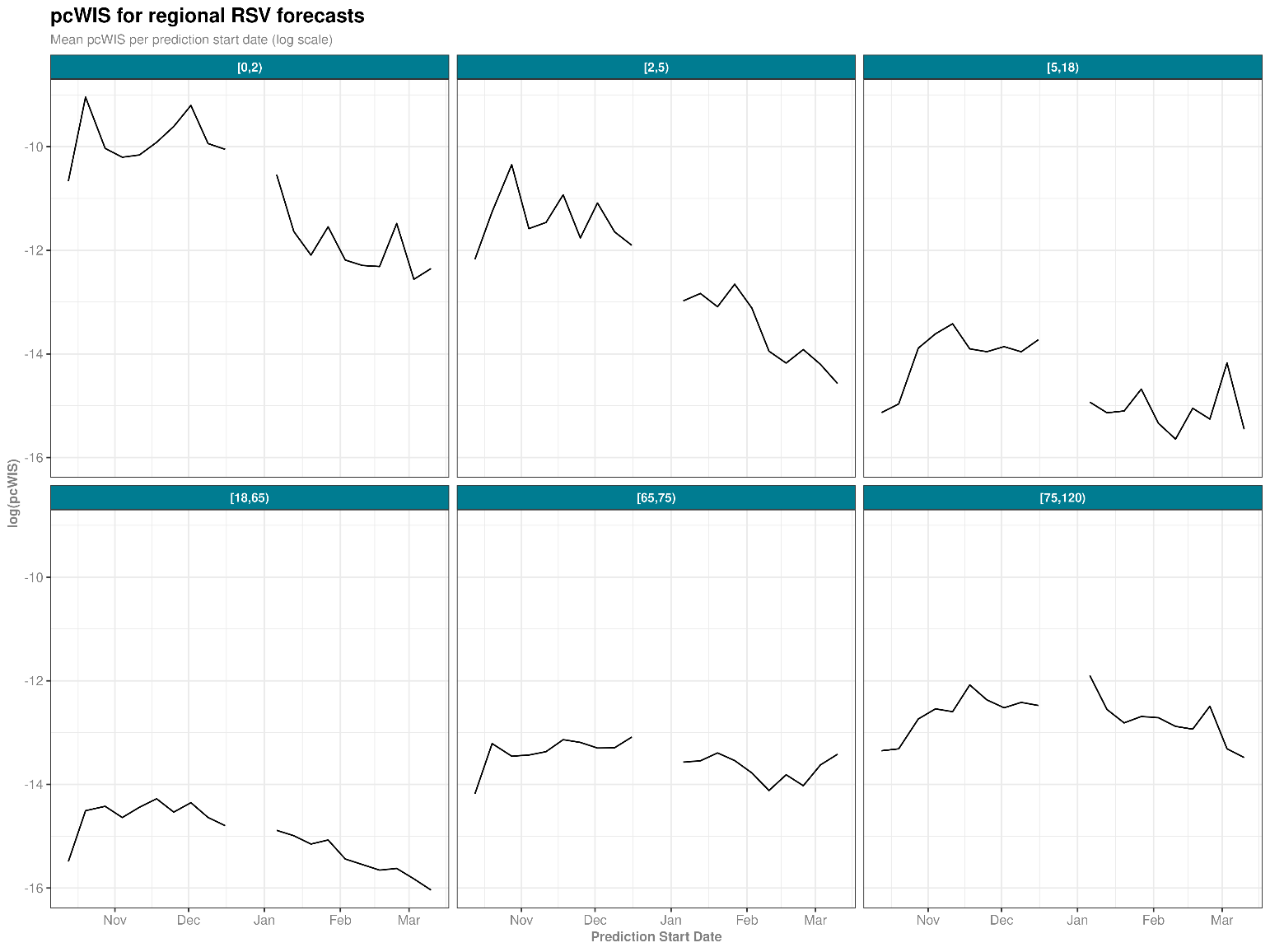

Figure G. log(pcWIS) by age group (national geography) and prediction start date for RSV operational ensemble forecasts. Performance is generally worse for the very young, or very old age groups who typically have higher incidence. Within age groups, forecast performance tends to increase slightly as we progress through the season. This could be attributed to (i) reduced case counts (ii) development of the ensemble in real time

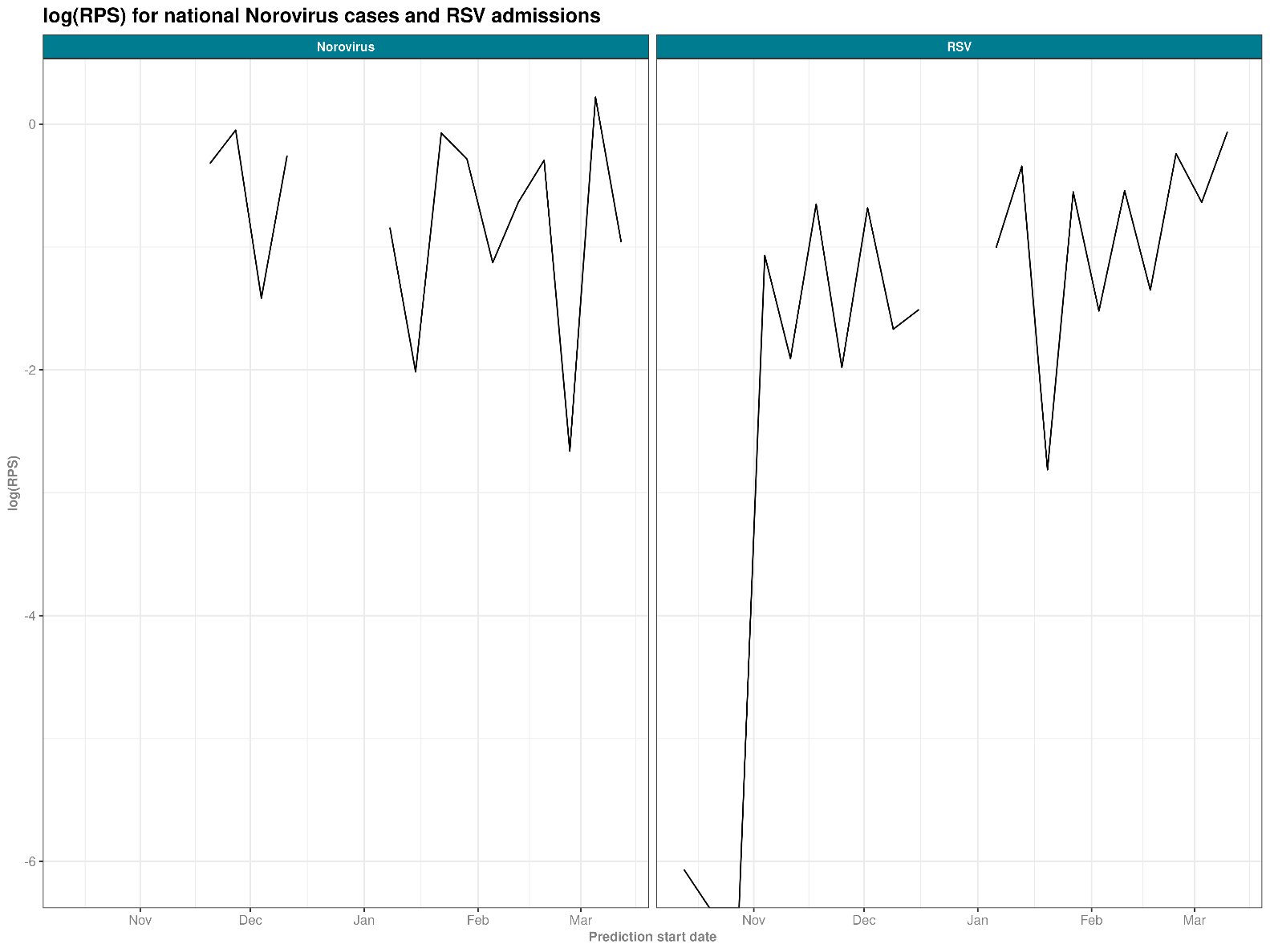

Figure H. log(RPS) by prediction start date for national single model Norovirus cases and national RSV operational ensemble forecasts. Norovirus scoring starts later in the season because (i) norovirus modelling started later in the season and (ii) trend direction estimation for norovirus was developed later in the season.

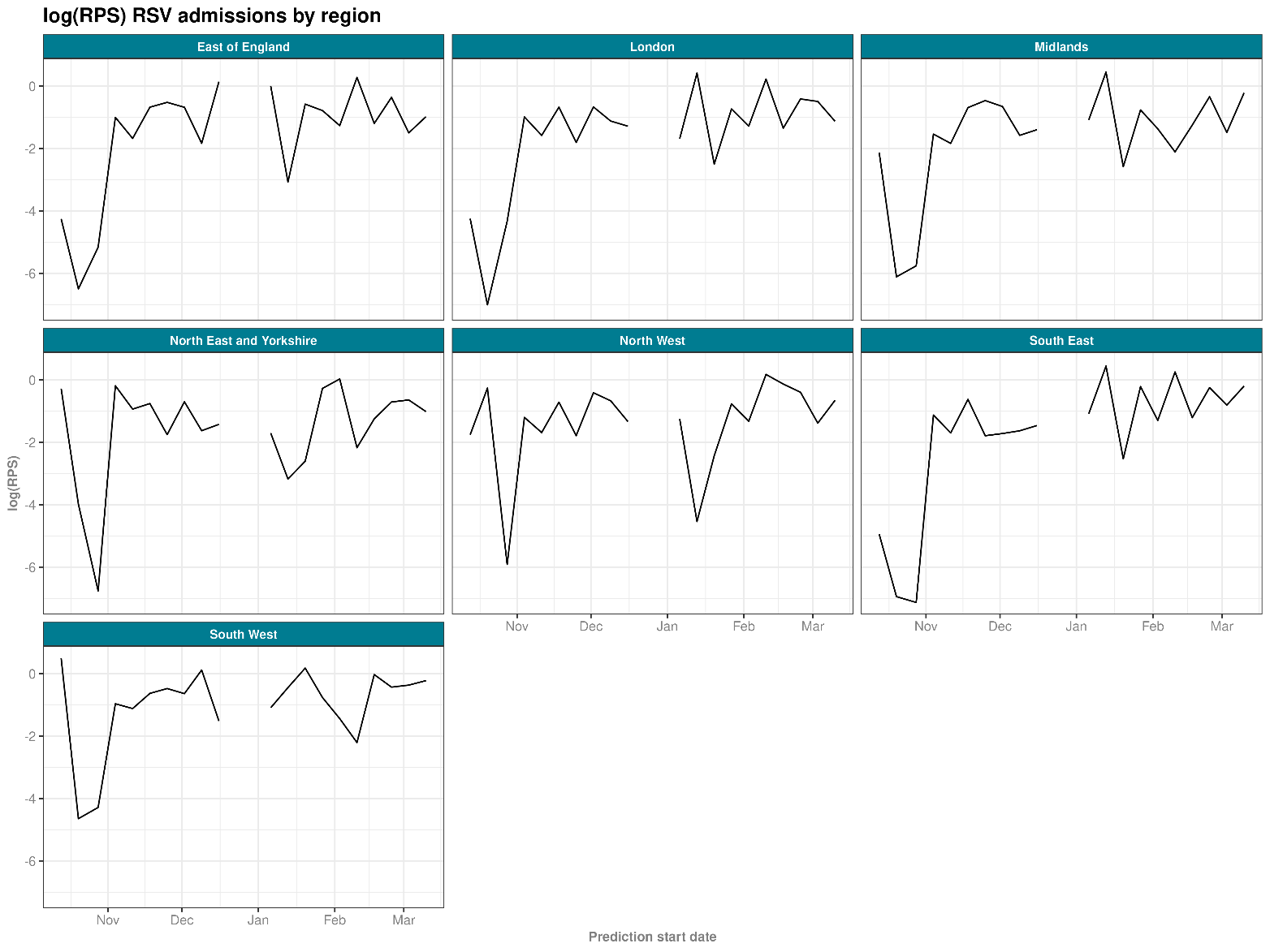

Figure I. log(RPS) by prediction start date and NHS Region for RSV operational ensemble admissions forecasts. Trend direction performance was similar across the season, with some early season estimates offering mixed results.

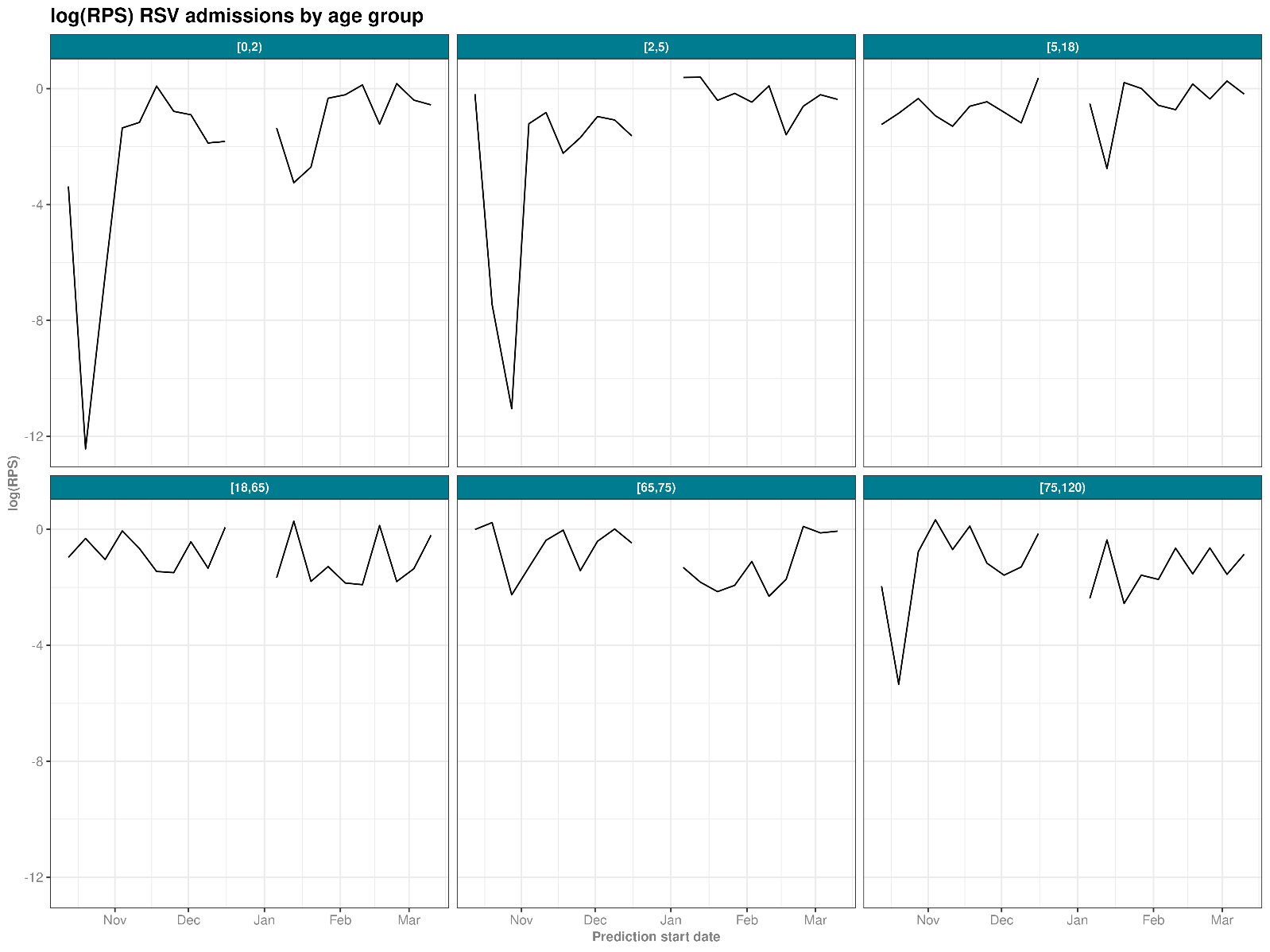

Figure J. log(RPS) by prediction start date and age group (national geography) for RSV operational ensemble admissions forecasts. Trend direction estimation performance was similar across the season, with some correct, and highly confident, forecasts early in the season in the two youngest age groups.

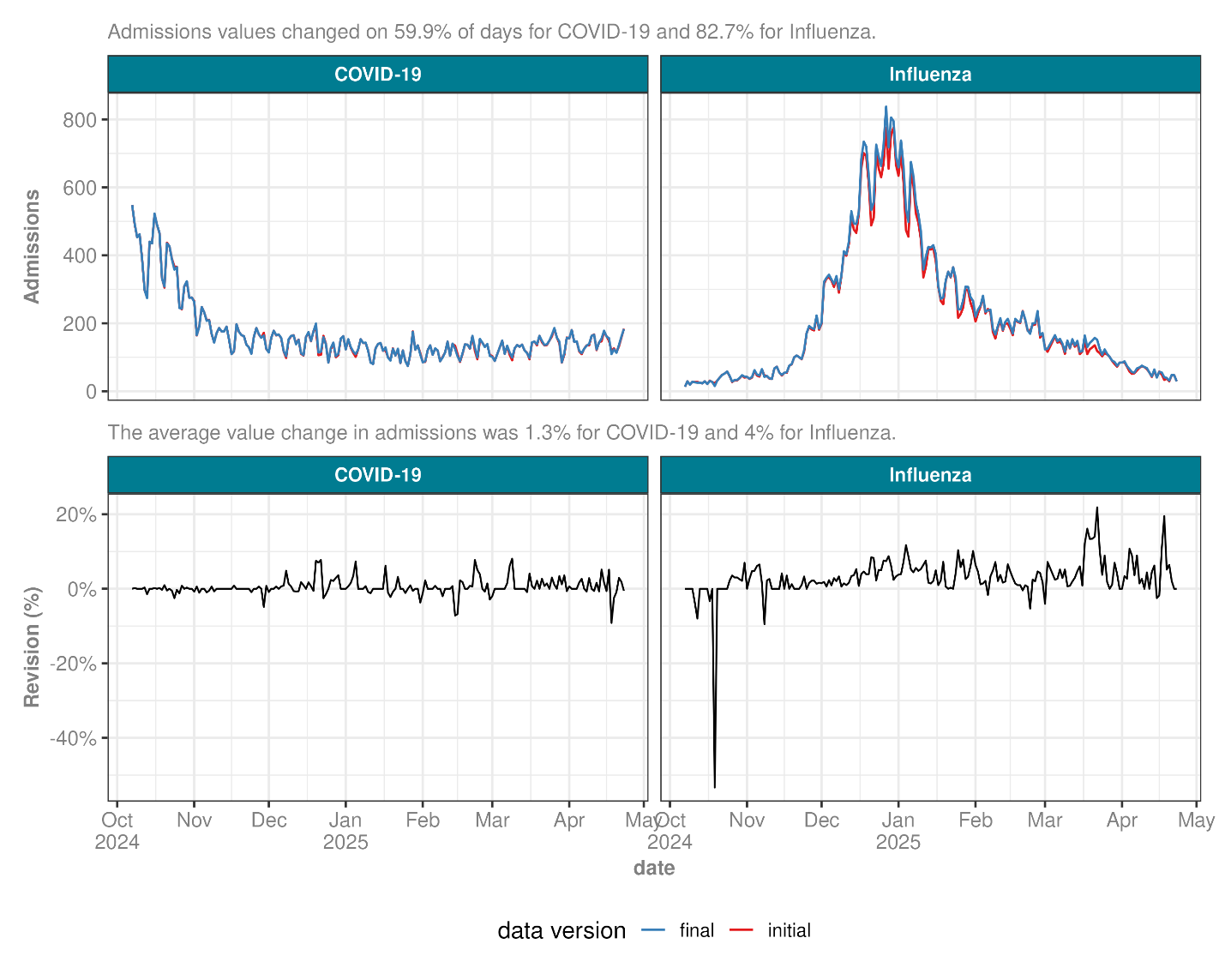

Figure K: (Top) A time series of admissions over time at the national level for COVID-19 and Influenza. The red line indicates the data UKHSA received initially, and in blue the final revised version of the data. (Bottom) The percentage change from the initially received admission count to the final counts over time for COVID-19 and Influenza. As national data is shown for simplicity, some local variation may be hidden due to cancelling directions. There are larger percentage changes for influenza at the start and end of the season where counts are low. The revisions for Influenza are largest near the winter holiday period, where operational forecasts were not reported.

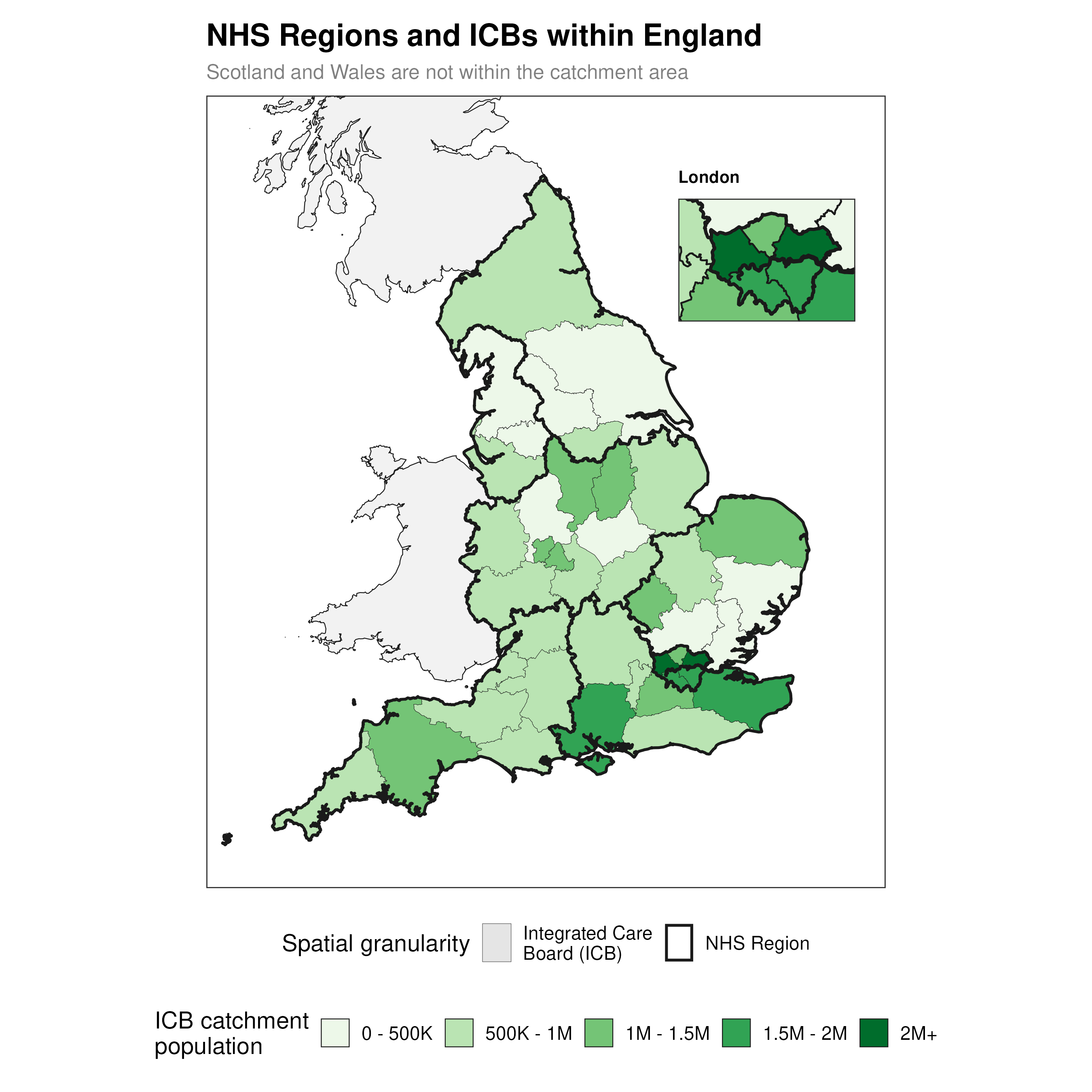

Figure L: Map illustrating NHS regions and ICBs in England, and their approximate population sizes. The smallest geographies are coloured by their population sizes. The NHS regions and noted by black bold lines. London, due to its relatively small geographical area, is shown in a cut-out in the top right of the figure. Data processed and figure generated using R, using spatial boundaries and population catchment data. Source: Office for National Statistics licensed under the Open Government Licence v.3.0. Contains OS data © Crown copyright and database right 2026. National boundaries: <https://geoportal.statistics.gov.uk/datasets/ons::countries-december-2024-boundaries-uk-buc-2/about> ; regional boundaries: <https://geoportal.statistics.gov.uk/datasets/ons::nhs-england-regions-january-2024-boundaries-en-bfc/about> ; ICB boundaries: <https://geoportal.statistics.gov.uk/datasets/ons::integrated-care-boards-april-2023-boundaries-en-bsc/about>

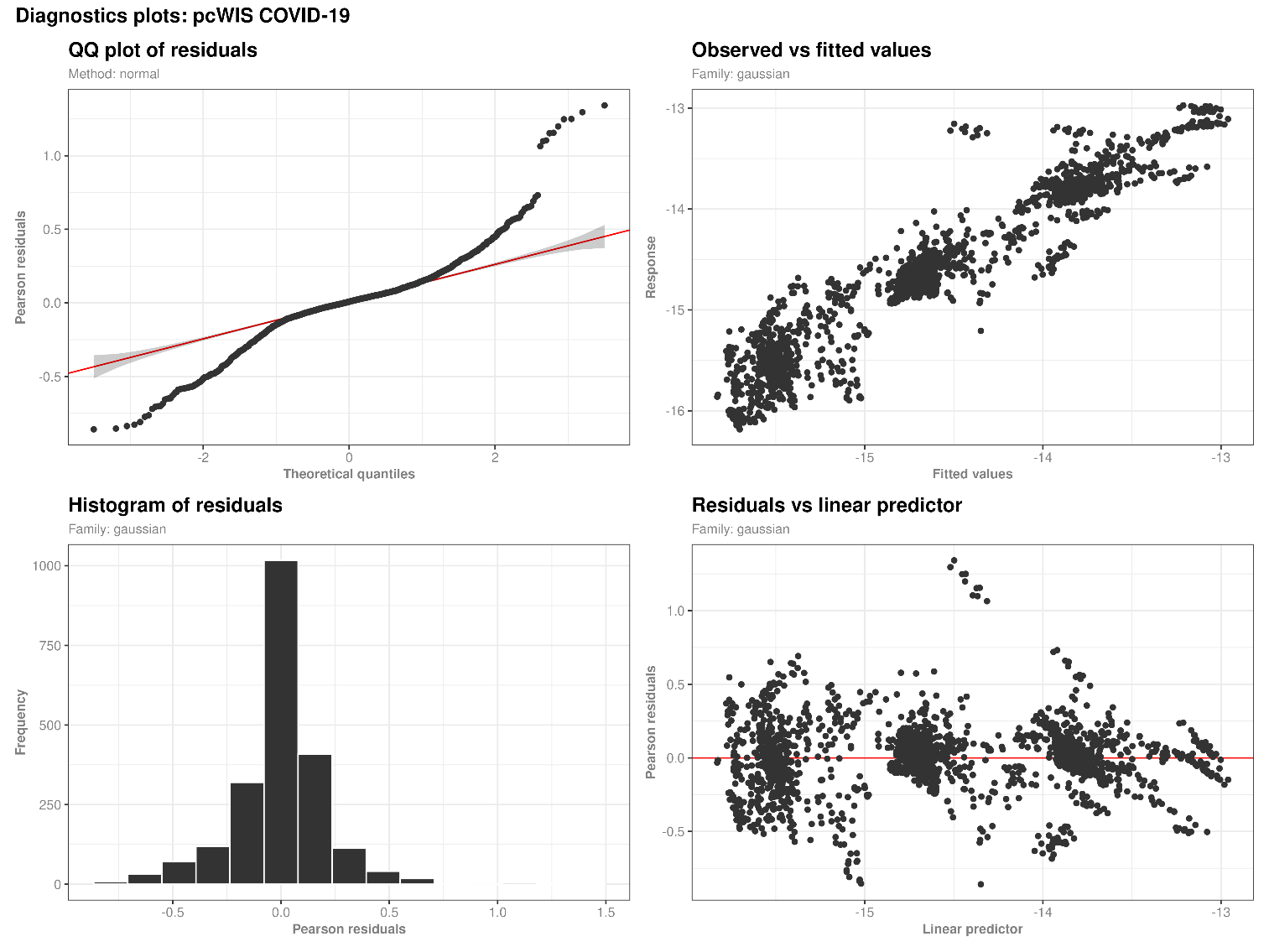

Figure M: Diagnostic plots for COVID-19 pcWIS GAM. QQ plot shows moderate deviation from Normality. Histogram shows approximately symmetric residuals and suggests heavy-tailed residuals. Observed vs fitted values shows an approximately linear relationship, with a small cluster of outliers. The Residuals vs linear predictor shows three main groups (one per location level) and a small cluster of potential outliers. Overall, there is moderate deviation from Normality and homoscedasticity of residuals which may affect inferential statements, but predicted values are reasonably close to observed values. This is expected to have only a minor impact on pcWIS analysis.

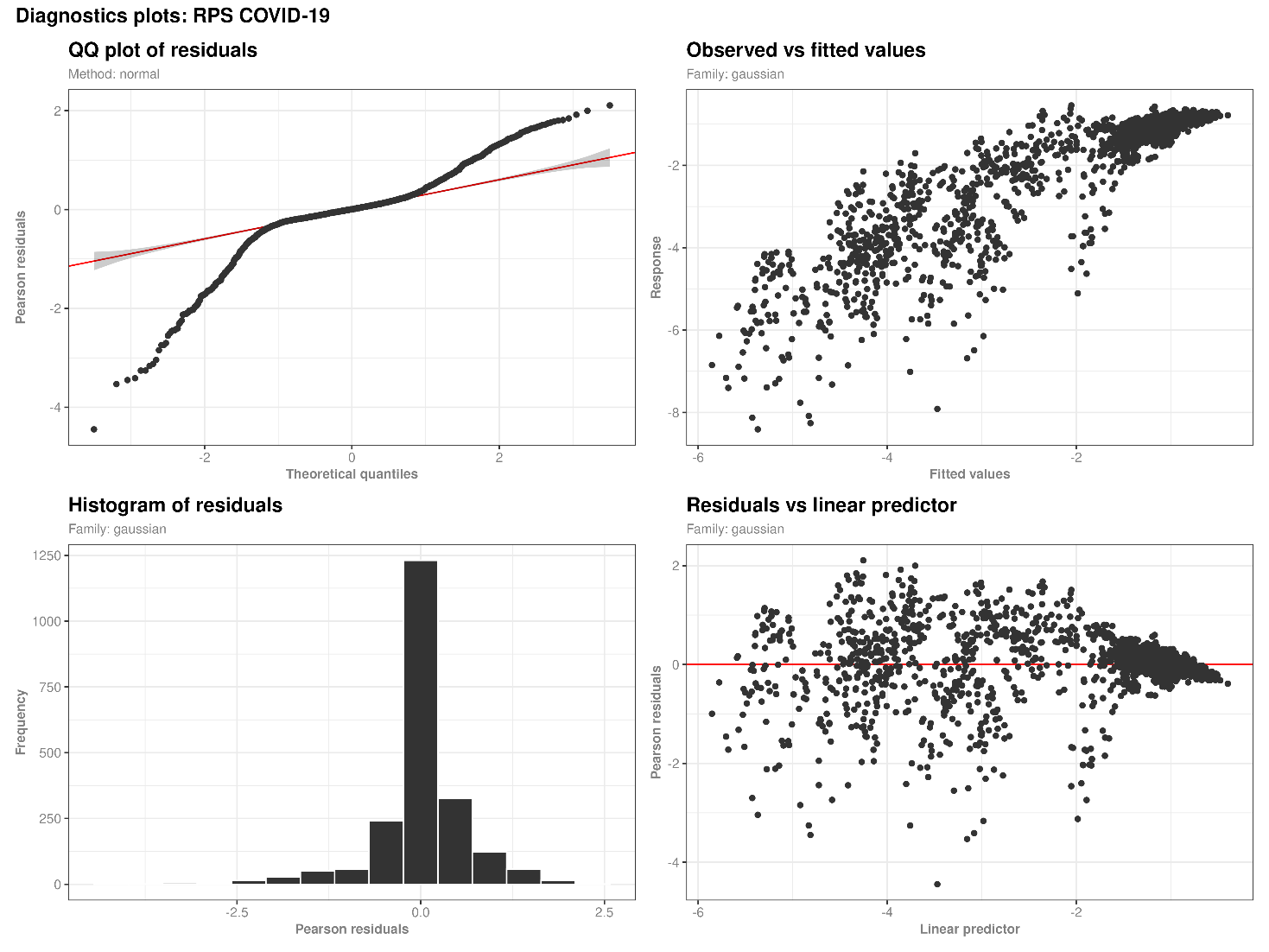

Figure N: Diagnostic plots for COVID-19 RPS GAM. QQ plot shows moderate deviation from Normality. Histogram shows approximately symmetric residuals, there is mild assymetry and heavy-tailed residuals. Observed vs fitted values shows an approximately linear relationship, there is some mild curvature and moderate scatter. The Residuals vs linear predictor shows a decrease in residual variability as the value of the linear predictor increases, but no other obvious patterns. Overall, there is moderate deviation from Normality and homoscedasticity of residuals which may affect inferential and predictive statements. This could have a moderate effect on inferences of the COVID-19 RPS analysis.

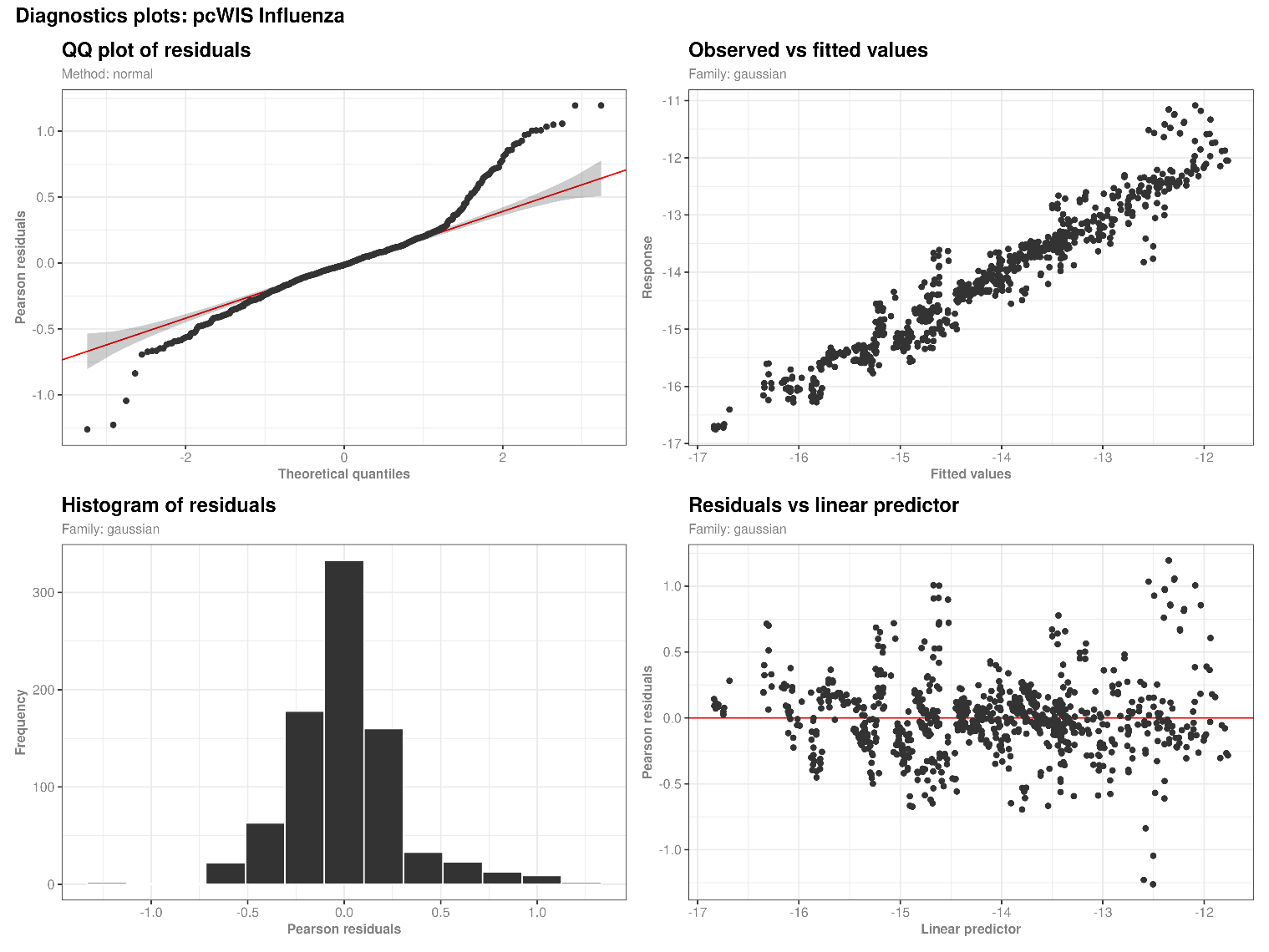

Figure O: Diagnostic plots for Influenza pcWIS GAM. QQ plot shows moderate deviation from Normality. Histogram shows mildly asymmetric residuals. Observed vs fitted values shows an approximately linear relationship, with a small group of potential outliers when the fitted values are large. The Residuals vs linear predictor some evidence of residual variance increasing with the linear predictor. Overall, there is mild deviation from Normality and homoscedasticity of residuals which may affect inferential statements, but predicted values are reasonably close to observed values. This is expected to have only a minor impact on Influenza pcWIS analysis.

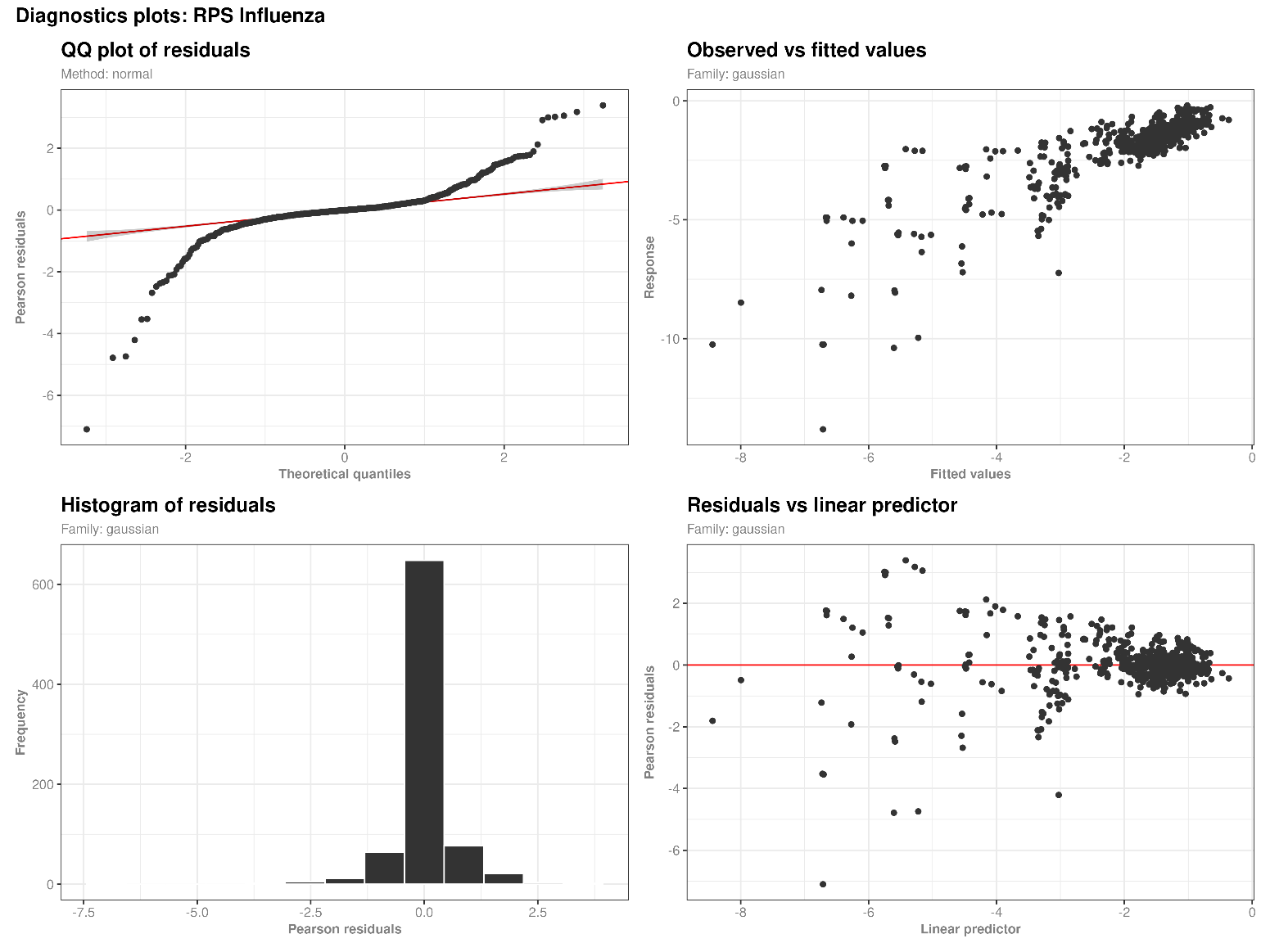

Figure P: Diagnostic plots for Influenza pcWIS RPS. QQ plot shows moderate deviation from Normality with one point having a very large, negative residual (approximate value, -6.5). Histogram shows mildly asymmetric residuals. Observed vs fitted values shows an approximately linear relationshipbut with variable scatter. The Residuals vs linear predictor some evidence of residual variance decreasing with the linear predictor. Overall, there is mild deviation from Normality and homoscedasticity of residuals which may affect inferential statements, but predicted values are reasonably close to observed values. The large potential outlier was for the gam_cr_gam_gp_ets sub-ensemble. This had a perfect RPS score of 0 which causes problems with a log transform. A small offset (${10}^{-6})$ was added to all RPS values in the Influenza analysis to avoid $\log\left( 0 \right)$. However, the GAM still struggled to adequately account for this value. Overall, this is expected to have a moderate impact on Influenza pcWIS analysis.

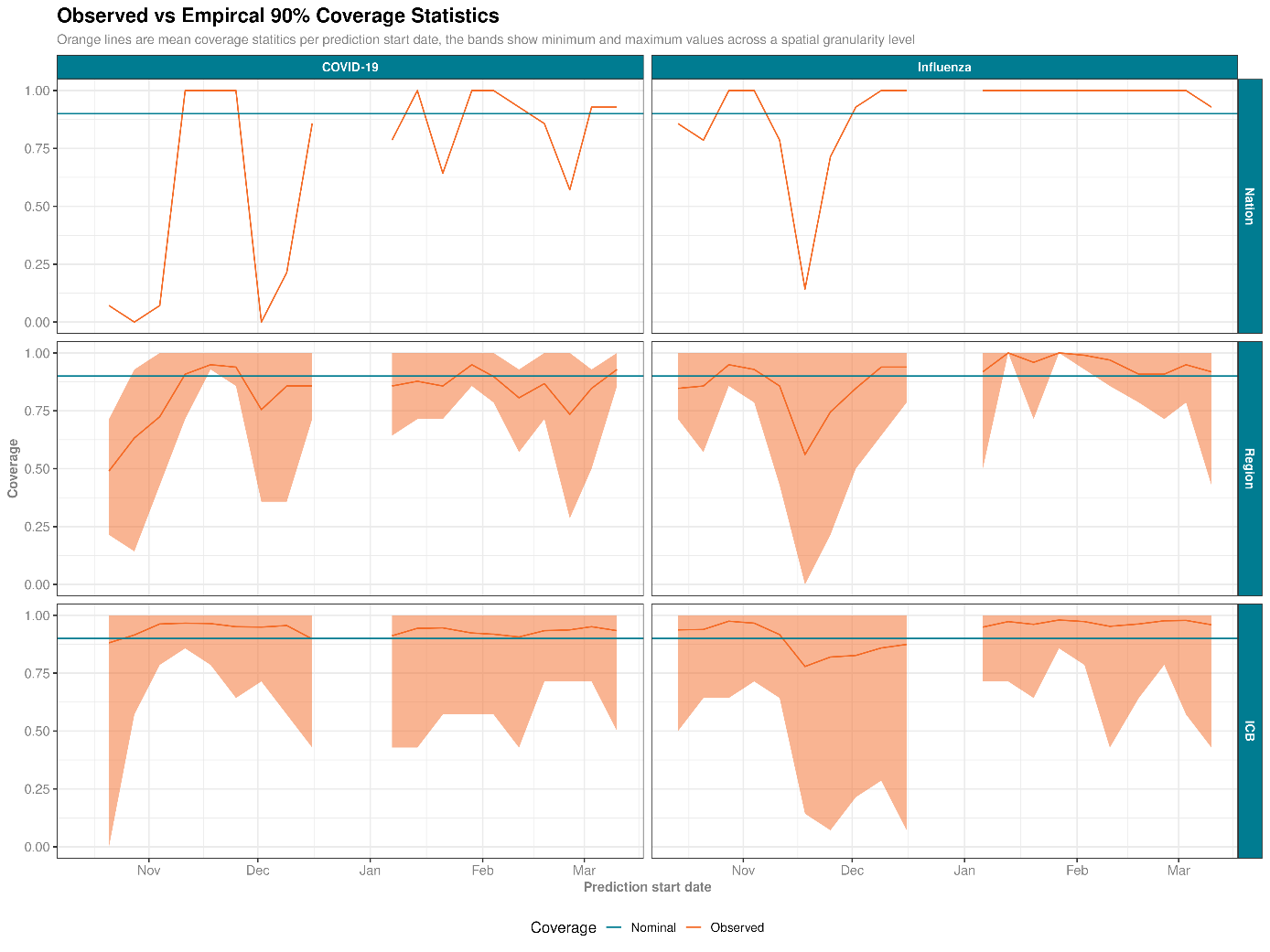

Figure Q: Comparison of observed and nominal coverage statistics for forecasts by disease and spatial granularity, averaged over the 14 day prediction horizon. No bands are present for the national forecasts as there is only one national forecast per prediction start date-disease combination. The orange bands for Region and ICB represent the min and max value present across the locations. The blue, horizontal line represents the nominal coverage (90%).

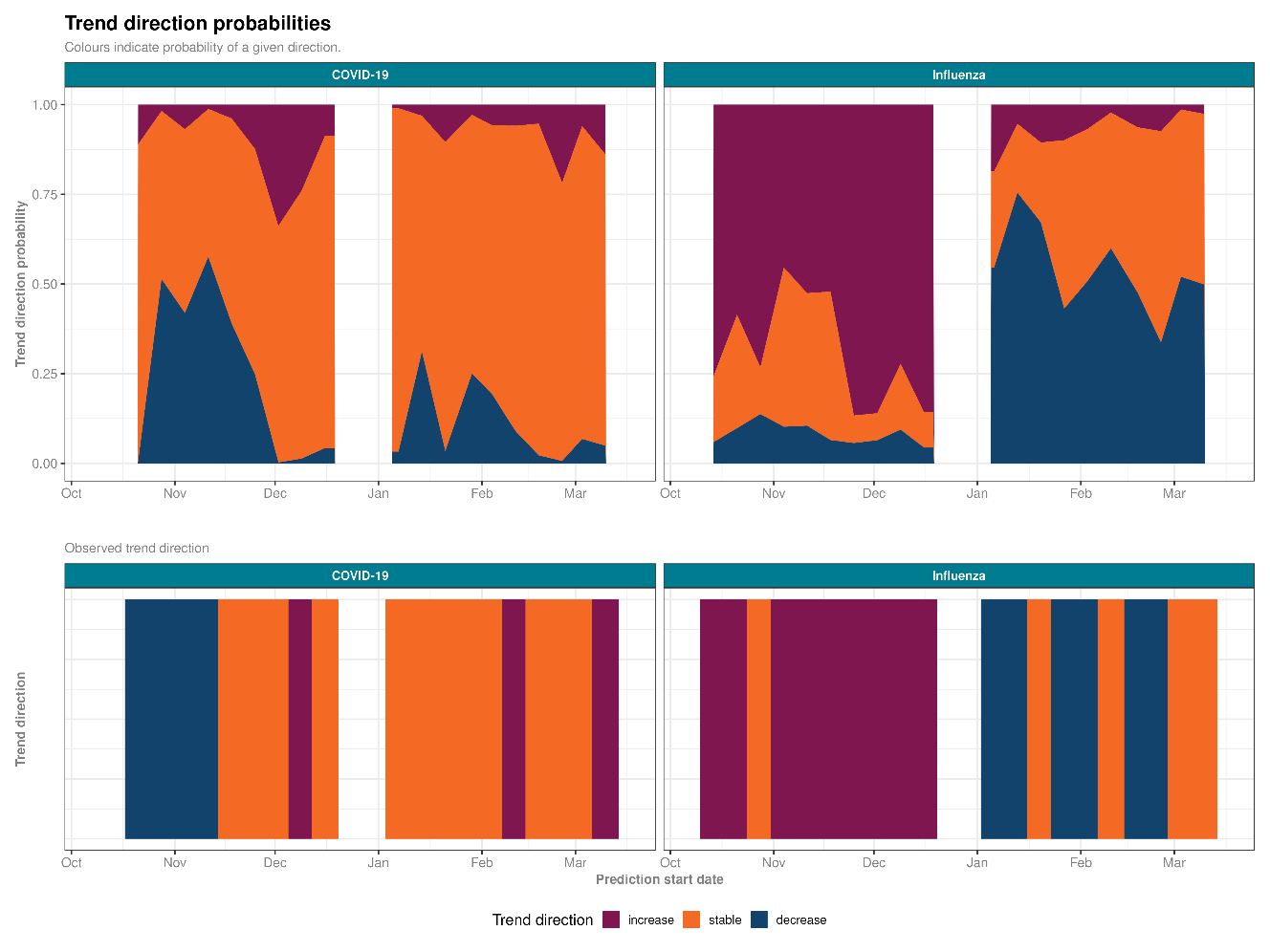

Figure R: (Top) Trend direction probability estimates for national COVID-19 and Influenza admissions forecasts of the operational ensemble at national geography. (Bottom) Observed epidemic trend directions at national geography.

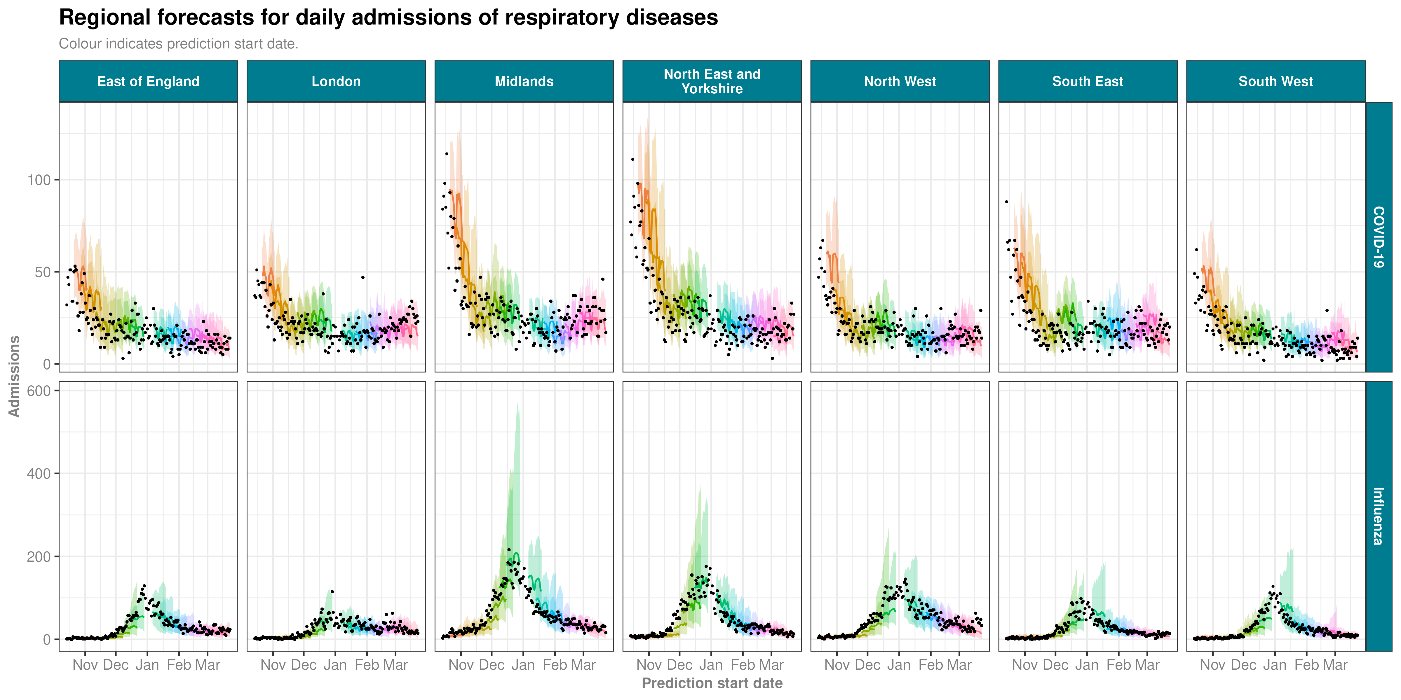

Figure S. Regional operational ensemble forecast for COVID-19 and Influenza. In these cases, the shapes of the admissions curves are similar to the corresponding national forecasts, but naturally, the number of admissions is lower at finer spatial resolutions.

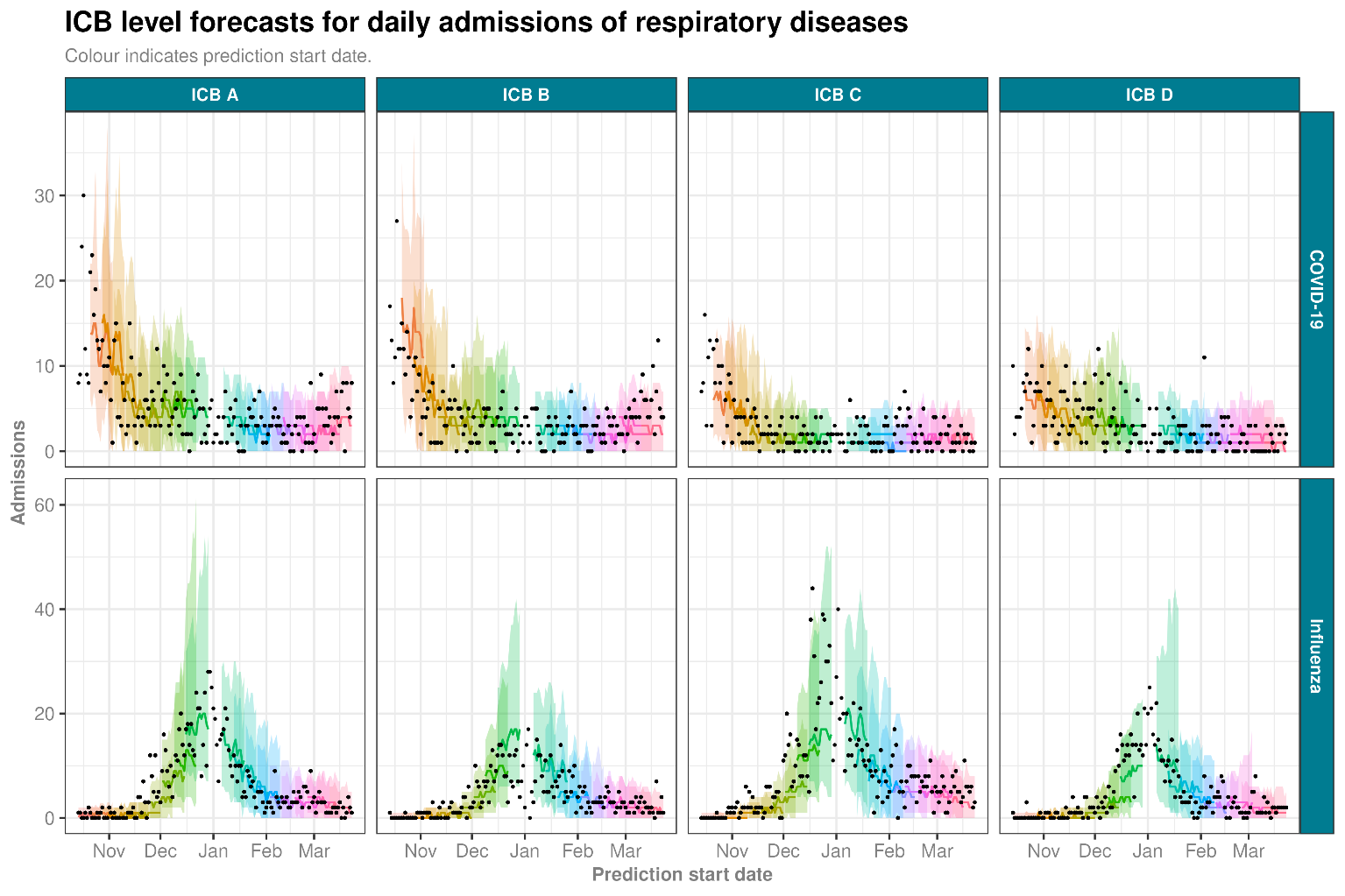

Figure T: Example ICB forecasts for daily COVID-19 and Influenza hospital admissions. ICB names are redacted for data governance reasons. Like regional forecasts, ICB forecasts tend to follow a similar epidemic curve to the corresponding national forecast, but the scale of the epidemic is much smaller.

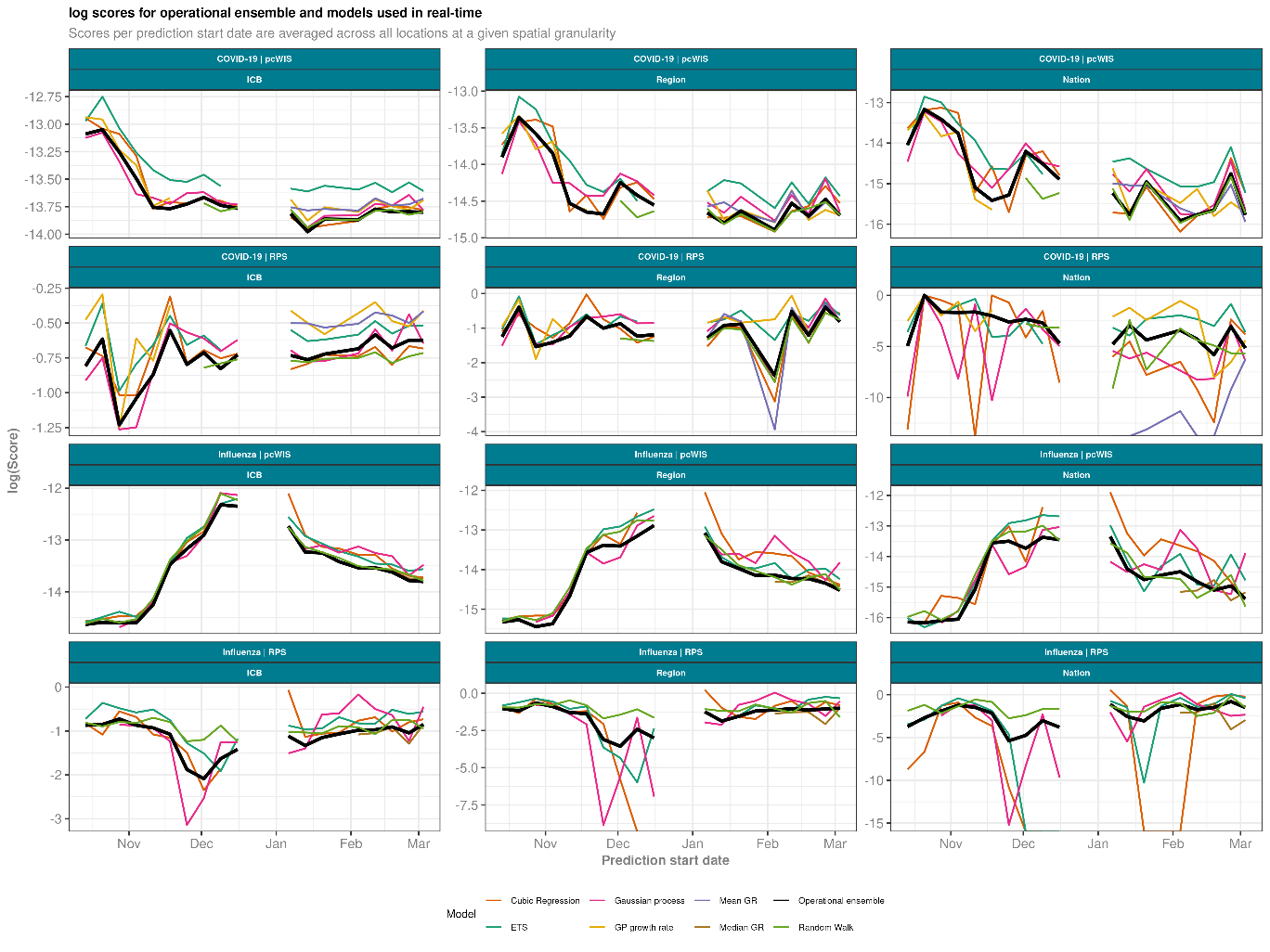

Figure U. log(pcWIS) and log(RPS) per disease-location level combination. Note for some individual models, the trend direction estiamte was near perfect, so the log scores diverge to $-\infty$ and are therefore not visible on the plot.

**Retrospective forecasts**

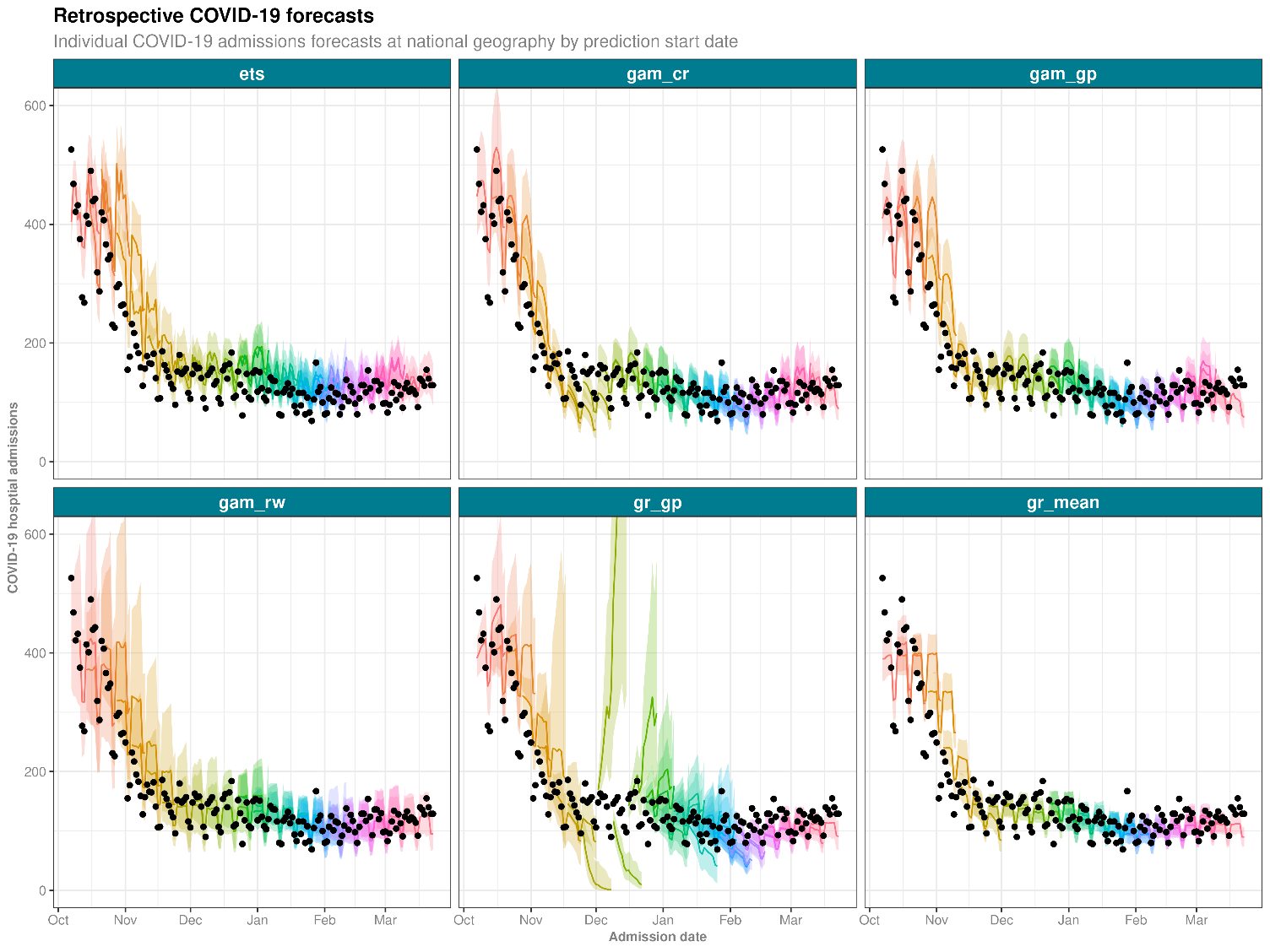

Figure V. Individual retrospective model predictions for COVID-19 hospital admission at national geography. The nation wave during the study period included a decline then stable period with limited variation.

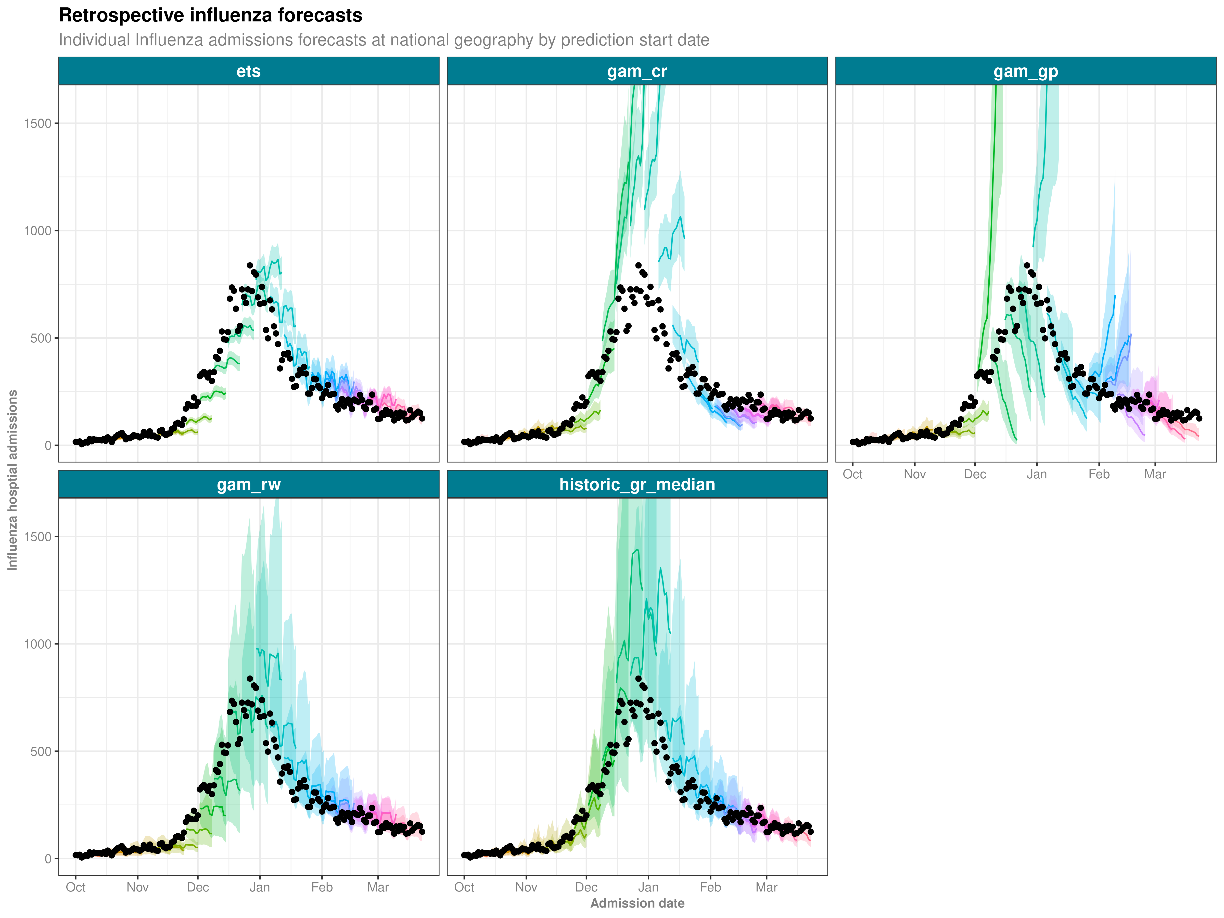

Figure W. individual retrospective model predictions for Influenza hospital admissions at national geography. The epidemic wave showed a sharp rise in admissions followed by a more gradual decline.

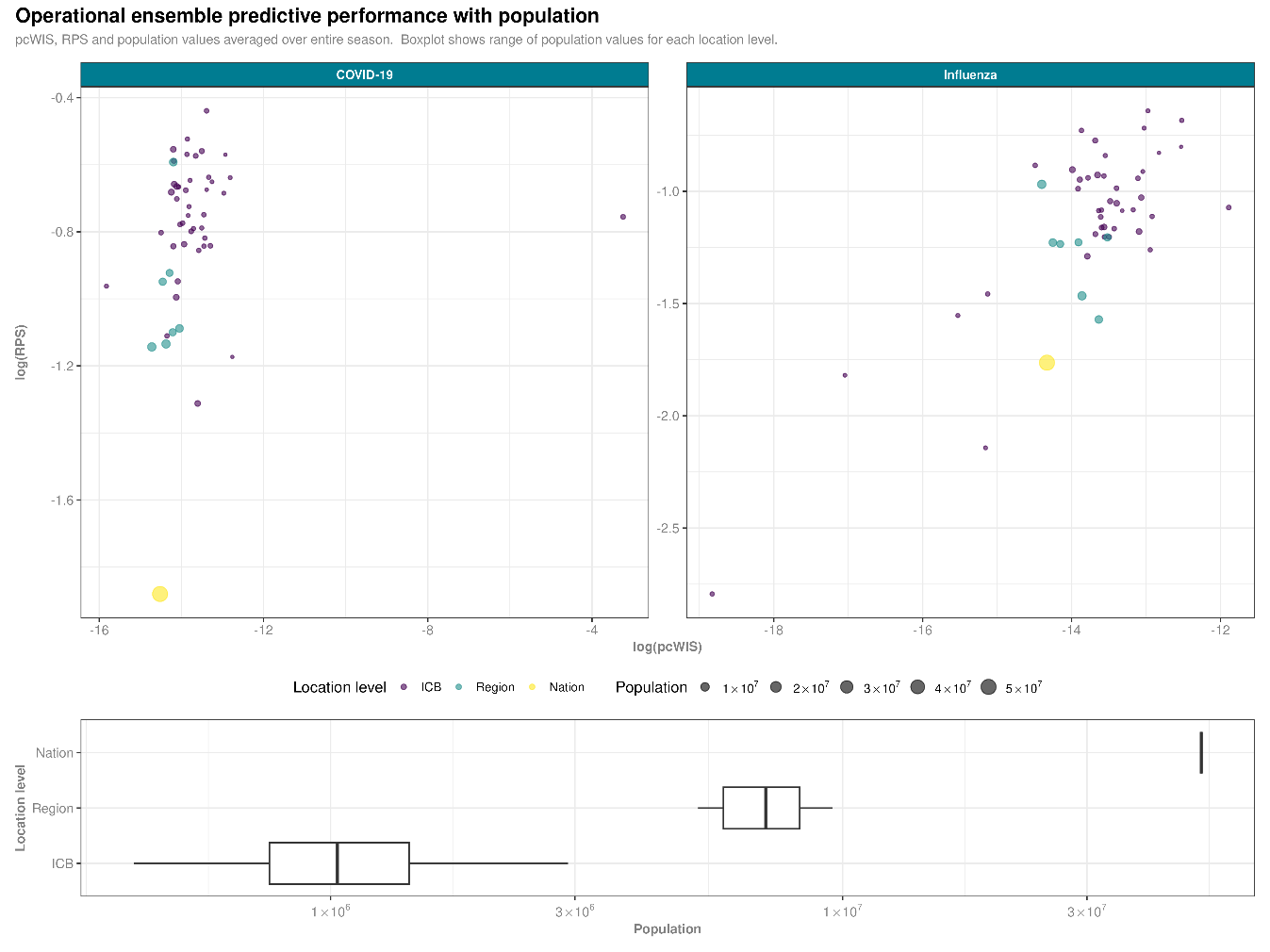

Figure X. (Top) Bubble chart showing log(RPS) vs log(pcWIS) for different forecasting locations, point size represents the population of the location for forecasting. (Bottom) Boxplot showing population sizes per location level. In general, scores are lower (better) for locations with a larger population.

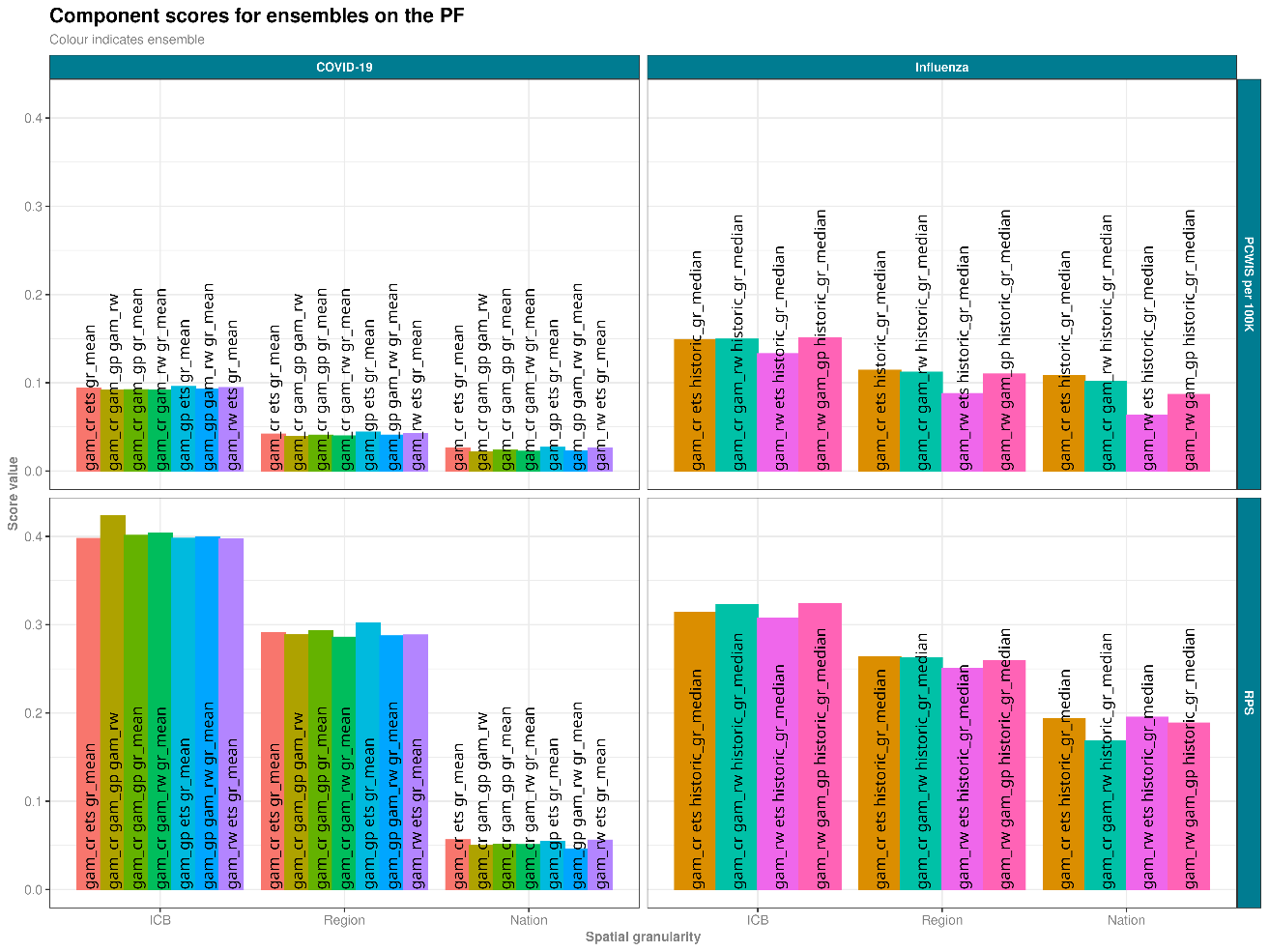

Figure Y. Values used in the Pareto front calculations (Figure 5).

| **Model Type** | **Disease** | **Score Name** | **Location Level** | **Mean Score** |
| --- | --- | --- | --- | --- |
| **COVID-19** |  |  |  |  |
| Individual Models | COVID-19 | pcWIS | Nation | $4.61 \times{10}^{-7}$ |
| Matched Ensemble | COVID-19 | pcWIS | Nation | $3.40 \times{10}^{-7}$ |
| Operational Ensemble | COVID-19 | pcWIS | Nation | $4.69 \times{10}^{-7}$ |
| Individual Models | COVID-19 | pcWIS | Region | $5.99 \times{10}^{-7}$ |
| Matched Ensemble | COVID-19 | pcWIS | Region | $5.26 \times{10}^{-7}$ |
| Operational Ensemble | COVID-19 | pcWIS | Region | $5.94 \times{10}^{-7}$ |
| Individual Models | COVID-19 | pcWIS | ICB | $1.52 \times{10}^{-3}$ |
| Matched Ensemble | COVID-19 | pcWIS | ICB | $1.11 \times{10}^{-6}$ |
| Operational Ensemble | COVID-19 | pcWIS | ICB | $9.84 \times{10}^{-4}$ |
| Individual Models | COVID-19 | RPS | Nation | $1.88 \times{10}^{-1}$ |
| Matched Ensemble | COVID-19 | RPS | Nation | $9.50 \times{10}^{-2}$ |
| Operational Ensemble | COVID-19 | RPS | Nation | $1.63 \times{10}^{-1}$ |
| Individual Models | COVID-19 | RPS | Region | $4.47 \times{10}^{-1}$ |
| Matched Ensemble | COVID-19 | RPS | Region | $2.93 \times{10}^{-1}$ |
| Operational Ensemble | COVID-19 | RPS | Region | $3.78 \times{10}^{-1}$ |
| Individual Models | COVID-19 | RPS | ICB | $5.37 \times{10}^{-1}$ |
| Matched Ensemble | COVID-19 | RPS | ICB | $3.96 \times{10}^{-1}$ |
| Operational Ensemble | COVID-19 | RPS | ICB | $4.85 \times{10}^{-1}$ |
| **Influenza** |  |  |  |  |
| Individual Models | Influenza | pcWIS | Nation | $1.34 \times{10}^{-6}$ |
| Matched Ensemble | Influenza | pcWIS | Nation | $6.75 \times{10}^{-7}$ |
| Operational Ensemble | Influenza | pcWIS | Nation | $5.62 \times{10}^{-7}$ |
| Individual Models | Influenza | pcWIS | Region | $1.54 \times{10}^{-6}$ |
| Matched Ensemble | Influenza | pcWIS | Region | $9.37 \times{10}^{-7}$ |
| Operational Ensemble | Influenza | pcWIS | Region | $8.55 \times{10}^{-7}$ |
| Individual Models | Influenza | pcWIS | ICB | $2.10 \times{10}^{-6}$ |
| Matched Ensemble | Influenza | pcWIS | ICB | $1.41 \times{10}^{-6}$ |
| Operational Ensemble | Influenza | pcWIS | ICB | $1.47 \times{10}^{-6}$ |
| Individual Models | Influenza | RPS | Nation | $2.71 \times{10}^{-1}$ |
| Matched Ensemble | Influenza | RPS | Nation | $1.60 \times{10}^{-1}$ |
| Operational Ensemble | Influenza | RPS | Nation | $1.61 \times{10}^{-1}$ |
| Individual Models | Influenza | RPS | Region | $4.15 \times{10}^{-1}$ |
| Matched Ensemble | Influenza | RPS | Region | $2.45 \times{10}^{-1}$ |
| Operational Ensemble | Influenza | RPS | Region | $2.88 \times{10}^{-1}$ |
| Individual Models | Influenza | RPS | ICB | $4.51 \times{10}^{-1}$ |
| Matched Ensemble | Influenza | RPS | ICB | $3.04 \times{10}^{-1}$ |
| Operational Ensemble | Influenza | RPS | ICB | $3.55 \times{10}^{-1}$ |

Supplementary Table 1. Mean scores for models used in winter operations. Smaller values indicate better forecasts. In all cases, the average ensemble score for each location level-disease-score group was less than the average operational individual models score.

| **Disease** | **Ensemble** |
| --- | --- |
| COVID-19 | gam_cr, ets, gr_mean |
|  | gam_cr, gam_gp, gam_rw |
|  | gam_cr, gam_gp, gr_mean |
|  | gam_cr, gam_rw, gr_mean |
|  | gam_gp, ets, gr_mean |
|  | gam_gp, gam_rw, gr_mean |
|  | gam_rw, ets, gr_mean |
| Influenza | gam_cr, ets, historic_gr_median |
|  | gam_cr, gam_rw, historic_gr_median |
|  | gam_rw, ets, historic_gr_median |
|  | gam_rw, gam_gp, historic_gr_median |

Supplementary Table 2. Members of Pareto Front for full Pareto analysis (not using weighted averages) by disease.

| **Disease** | **Spatial granularity** | **Observed 90% coverage** | **Observed 50% coverage** |
| --- | --- | --- | --- |
| COVID-19 | Nation | 87.5% | 56.3% |
|  | Region | 91.2% | 53.7% |
|  | ICB | 95.2% | 65.5% |
| Influenza | Nation | 90.6% | 62.0% |
|  | Region | 91.8% | 59.9% |
|  | ICB | 94.6% | 71.0% |

Supplementary Table 3. Comparison of nominal coverage of prediction intervals to observed coverage by disease and spatial granularity for operational ensembles. Statistics are an average for the entire season and where relevant, averaged across locations.

| **Disease** | **Metric** | **Minimum** | **Overall mean** | **Maximum** | **Operational mean** |
| --- | --- | --- | --- | --- | --- |
| COVID-19 | pcWIS | $2.03\times{10}^{-7}$ | $2.39\times{10}^{-6}$ | $3.27\times{10}^{-7}$ | $3.98\times{10}^{-7}$ |
| COVID-19 | RPS | $4.53\times{10}^{-2}$ | $2.69\times{10}^{-2}$ | $8.60\times{10}^{-2}$ | $2.42\times{10}^{-1}$ |
| Influenza | pcWIS | $6.69\times{10}^{-7}$ | $1.06\times{10}^{-6}$ | $1.31\times{10}^{-6}$ | $5.20\times{10}^{-7}$ |
| Influenza | RPS | $1.64\times{10}^{-1}$ | $1.90\times{10}^{-1}$ | $2.14\times{10}^{-1}$ | $2.34\times{10}^{-1}$ |

Supplementary Table 4. Minimum, overall mean, and maximum values of retrospective ensemble scores averaged across the season for national predictions; with operational ensemble average for comparison.

| **Model type** | **Location level** | **Age group granularity** | **Disease** | **Score name** | **Mean score** |
| --- | --- | --- | --- | --- | --- |
| **Norovirus** |  |  |  |  |  |
| Single operational model | Nation | None | Norovirus | pcWIS | $2.20 \times{10}^{1}$ |
| Single operational model | Nation | None | Norovirus | RPS | $5.88 \times{10}^{-1}$ |
| **RSV** |  |  |  |  |  |
| Operational ensemble | Nation | None | RSV | pcWIS | $1.04 \times{10}^{-6}$ |
| Operational ensemble | Nation | Fine | RSV | pcWIS | $5.98 \times{10}^{-6}$ |
| Operational ensemble | Region | None | RSV | pcWIS | $1.55 \times{10}^{-6}$ |
| Operational ensemble | Nation | None | RSV | RPS | $4.56 \times{10}^{-1}$ |
| Operational ensemble | Nation | Fine | RSV | RPS | $5.83 \times{10}^{-1}$ |
| Operational ensemble | Region | None | RSV | RPS | $5.01 \times{10}^{-1}$ |

Supplementary Table 5. Mean scores for models used in winter operations. Smaller values indicate better forecasts. In all cases.

### Supplementary Section B

A specification of the novel models added in the 2024-2025 season for COVID-19 and Influenza forecasts. Hyperparameters are provided within the configuration files in <https://github.com/jcken95/sub-ensemble-evaluation>.

GP growth rate model (COVID-19)

At a high level the aim of this model was to estimate a crude instantaneous growth rate from hospital admissions, then forecast the GR forward in time using a Gaussian process. Using this forecasted growth rate and recent values of admissions, future admissions could be simulated.

At a high level the model:

- calculates a local growth rate based on rolling averages of the target

- estimate this local growth rate with a short training length and Gaussian process model in time

- apply this forecasted growth rate forward in time to the most recent target.

Throughout we will refer to the growth rate as $GR_{t}=\log\left( s\left( \frac{Y_{t}}{Y_{t-1}} \right) \right)$

Where $s\left( \right)$ is some smoothing function - in our case a seven day rolling mean and $Y_{1}, Y_{2}\ldots Y_{t}$ is our admissions time series.

We then define our regression as

$$\mathrm{GR}_{t} = \mathrm{icb} + GP_{\mathrm{region}}\left( t \right)+ GP_{\mathrm{nation}}\left( t \right)$$

$\mathrm{icb}$ is a random effect for the ICB, $\mathrm{GP}_{location level}$ is a Gaussian process smooth pooled at the given location level. Once the $\mathrm{GR}$ has been estimated we need to convert back to admissions.

We take the forecasted growth rate up to $\mathrm{GR}_{t_{\max}+14}.$ The smooth estimates of admissions $s\left( Y_{t_{\max}} \right)$ are taken which can be extrapolated from.

The forecast for $t>t_{max}$ is therefore the summed growth rate up to $t$.

$$s\left( Y_{t} \right)=s\left( Y_{t_{\max}} \right)\times e^{\sum\mathrm{GR}_{t}}$$

However, this gives the smoothed admissions, so we can incorporate uncertainty according to a $\mathrm{NegBin}$.

$$Y_{t} \sim\mathrm{NegBin}\left( s\left( y_{t}), \theta\right) \right)$$

The model is tuned primarily by:

- how much data is fed into the model (training length)

- varying GP hyperparameters

- how long the rolling averages on the model smoothers are.

- dispersion parameter $\theta$.

Mean GR model (COVID-19)

The premise of this model is similar to that of “GP growth rate”, however, instead of a Gaussian process to estimate the time varying instantaneous growth, we estimate a short term fixed growth rate for each ICB. This fixed growth rate is then assumed to continue forward in time. Similarly, the future growth rate is then applied to recent admissions values to simulate future growth.

At a high level the model:

- calculates a local growth rate based on rolling averages of the target

- estimate this local growth rate with a short training length and linear model in time

- apply this forecasted growth rate forward in time to the most recent target.

Throughout we will refer to the growth rate as $GR_{t}=\log\left( s\left( \frac{Y_{t}}{Y_{t-1}} \right) \right)$

Where $s\left( \right)$ is some smoothing function - in our case a seven day rolling mean and $Y_{1}, Y_{2}\ldots Y_{t}$ is our admissions time series.

We then define our regression as

$$\mathrm{GR}_{t} = \mathrm{icb}_{\mathrm{region}}$$

$\mathrm{icb}_{\mathrm{region}}$ is a nested random effect for the ICB in each region. Once the $\mathrm{GR}$ has been estimated we need to convert back to admissions.

We take the forecasted growth rate up to $\mathrm{GR}_{t_{\max}+14}.$ The smooth estimates of admissions $s\left( Y_{t_{\max}} \right)$ are taken which can be extrapolated from.

The forecast for $t>t_{max}$ is therefore the summed growth rate up to $t$.

$$s\left( Y_{t} \right)=s\left( Y_{t_{\max}} \right)\times e^{\sum\mathrm{GR}_{t}}$$

However, this gives smoothed admissions, so we can incorporate uncertainty according to a $\mathrm{NegBin}$.

$$Y_{t} \sim\mathrm{NegBin}\left( s\left( y_{t}), \theta\right) \right)$$

The model is tuned primarily by:

- how much data is fed into the model (training length)

- how long the rolling averages on the model smoothers are.

- dispersion parameter $\theta$.

Median GR model (Influenza)

Conceptually the aim for this model is to leverage, in a straightforward way, past trends of influenza admissions. Using the SARI-Watch surveillance system, a weekly growth rate for each of the past seasons is calculated. While Influenza is highly heterogeneous each season, there are some generalized patterns each year, particularly in terms of direction rather than magnitude. A median growth rate from past seasons is estimated. Matching the current admissions data to past epiweeks we know the next two weeks’ median growth rate, which is applied to the current admissions data to produce a forecast. To generate uncertainty and day-of-week effects, we fit a random walk model to the data assuming a national trend, then apply the growth rate correction to this random walk forecast.

The core assumption is that there is information in past season growth rates that can help predict this season. This would be particularly helpful to capture early and late season dynamics, as well as behavioral changes that occur due to times of year (such as holiday periods).

The assumption is not about absolute values, but rather relative changes over time. Past seasons are matched to the current one by epidemic week `lubridate::epiweek`. To incorporate this trend we use a GAM RW model first, of the form:

$$Y_{t}\sim\mathrm{NegBin}\left( \mu_{t}, \theta\right)$$

$$\log E\left( Y_{t} \right)= \mathrm{do}w_{t} +\mathrm{ic}b_{\mathrm{region}} + W_{t} +\log\left( \mathrm{population} \right)$$

Where $W_{t}$ is a random walk; if $Z_{\tau}\sim N\left( 0, \sigma\right)$ are i.i.d. random variables then:

$W_{t}=\sum_{\tau=1}^{t} Z_{\tau}$ is a Gaussian random walk.

$Y_{t}$ is the number of admissions at time $t$, $\mathrm{do}w_{t}$ is a fixed effect for the day of week, $\mathrm{ic}b_{\mathrm{region}}$ is a random effect by ICB, nested within regions and $\log\left( \mathrm{population} \right)$serves as an offset term.

With this model, we take the past observed growth rates (in this case, the median of previous seasons). We accumulate the historic daily growth rate $gr_{t}$ over the prediction horizon as $GR_{t}$. For $t\geq prediction start date, Y_{t}=Y_{t}*GR_{t}$. Which gives a multiplicative factor based on past data and a random walk.

For the 24/25 season, the following past Influenza seasons were used to estimate the median: 2016-17, 2017-2018, 2018-2019, 2022-2023, 2023-2024. These years represent a mixture of pre-and-post COVID-19 public health social measures years.
